# Disability-relevant demyelination in multiple sclerosis: five-year evolution following diagnosis

**DOI:** 10.64898/2026.11.05.26367270

**Authors:** Elizabeth N. York, Rozanna Meijboom, Michael J. Thrippleton, Mark E. Bastin, Agniete Kampaite, Sarah-Jane Martin, Patrick K. A. Kearns, David P.J. Hunt, Siddharthan Chandran, Adam D. Waldman, the FutureMS consortium

**Author notes:** Correspondence to: Elizabeth N. York Centre for Clinical Brain Sciences, University of Edinburgh Edinburgh BioQuarter: Chancellors Building Edinburgh EH16 4SB, United Kingdom.

## Abstract

Improved understanding of how myelin damage contributes to disability in multiple sclerosis (MS) is needed to inform targeted therapeutic strategies and improve clinical trial platforms. Magnetization transfer saturation (*MTsat*) and *T*_1_ relaxometry provide *in vivo* quantitative MRI measures of myelin integrity, and may offer insights into disability-relevant pathology beyond inflammatory relapse. This longitudinal study explores the relationship between early demyelination and disability progression over the first five years following diagnosis.

62 people with relapsing-remitting (pwRR)MS (≤ 6 months post-diagnosis, 48 female, mean age 38 years) underwent baseline and one-year MRI and Expanded Disability Status Scale (EDSS) assessment. At five-year follow-up, 56/62 (90.3%) completed EDSS and 40/62 additionally completed MRI (64.5%). Disability progression was defined as an increase of ≥ 1.5, ≥ 1.0 or ≥ 0.5 points for a previous EDSS score of < 1.0, 1.0 to 5.5, and ≥ 6.0, respectively. Baseline neurofilament light chain (NfL), glial fibrillary acidic protein (GFAP), and EBNA1 immunoglobulin (IgG) titres were also measured. *MTsat* was calculated within normal-appearing white matter (NAWM), white matter lesions (WML), cortical and deep grey matter (GM) structures. Associations between change in *MTsat* and disability progression were assessed with binomial logistic regression. Interactions with interval disease activity (i.e. new/enlarging lesions [NELs]) were investigated, and comparisons were made with healthy control test-retest data (n = 11). Exploratory analyses were performed for: (prolonged) *T*_1_; baseline metrics; blood-derived markers; diffusion-weighted imaging; and, disability-associated differences in five-year microstructural MRI trajectories.

Greater baseline-to-one-year decreases in NAWM and WML *MTsat* were associated with baseline-to-one-year disability progression (adjusted odds ratio [95% confidence interval] = 0.255 [0.078 to 0.602] and 0.261 [0.084 to 0.638]; false discovery rate [FDR]-corrected *p*-value = 0.015 and 0.015). NELs did not influence these associations (interaction *p*-values > 0.05). One-to-five-year changes in *MTsat* were not associated with one-to-five-year disability progression (FDR-corrected *p*-values > 0.05). Neither baseline nor baseline-to-one-year *MTsat* changes predicted future disability progression. No exploratory analyses reached FDR-corrected statistical significance.

We show for the first time that progressive NAWM and WML demyelination measured with MTsat is associated with early disability progression in recently diagnosed RRMS, independent of radiological inflammatory disease activity. Neither *MTsat* nor *T*_1_ are associated with longer-term progression over the subsequent four years. MTsat is sensitive to disability-relevant demyelination not visible on conventional MRI, indicates a potential early window of therapeutic opportunity in the year following diagnosis, and is a promising clinically applicable disease stratifier and surrogate endpoint for therapeutic trials.

## Introduction

Immune-mediated demyelination in relapsing-remitting multiple sclerosis (RRMS) is thought to result in axonal damage, leading to neuronal dysfunction and clinical disability.^1,2^ Modulation of neuroinflammatory pathways with current disease-modifying therapies (DMTs) reduces rates of clinical relapse and new or enlarging white matter lesions (NELs), and slows disability progression.^3^ It is increasingly recognised however that disability accumulation in RRMS may also occur independently of DMT-modifiable inflammatory relapse activity, termed ‘silent progression’ or progression independent of relapse activity (PIRA).^4^ The pathological processes underlying disability progression in early RRMS and the variable trajectories of evolving disability for individual patients, however, remain poorly understood.

Current clinical neuroimaging criteria provide only limited insights into the biological processes that may be driving disability in early RRMS: the presence of NELs on serial T_2_- dependent MRI is an indicator of inflammatory relapse-associated radiological interval disease activity,^5^ and contrast-enhancing lesions (CELs) reflect blood-brain barrier breakdown, which is a surrogate marker of active inflammation. White matter lesion (WML) burden does not correlate well with clinical disability progression in RRMS, which has given rise to the widely-recognised notion of the ‘clinico-radiological paradox’.^6,7^ Grey matter (GM) lesions appear more closely associated with disability, but are more difficult to detect and quantify reliably on MRI.^8^

Emerging therapeutic strategies, focused on remyelination and neuroprotection, require more specific, objective, non-invasive *in vivo* measures of disability-relevant myelin and axonal damage for patient stratification, to assess target engagement, and as surrogate endpoints of clinical efficacy. Brain atrophy measured from longitudinal analysis of structural MRI has recently been proposed as an *in vivo* marker of neurodegeneration for clinical evaluation and therapeutic trial endpoints in MS.^9^ Widespread atrophy is apparent in early MS,^10^ has been shown to be reduced by DMTs,^11^ and is also greater in people with RRMS with PIRA than those who are clinically stable.^12^ Macrostructural atrophy however remains an indirect ‘downstream’ marker that may be confounded by other factors such as pseudoatrophy and brain hydration state,^9^ and lacks specificity for underlying pathophysiological features in brain tissue.

Diffuse myelin and axonal damage within normal-appearing white matter (NAWM) and grey matter (NAGM) are known pathological features of RRMS,^13^ and microstructural MRI allows more specific non-invasive quantification of myelin and axonal integrity than is possible with conventional structural imaging.^14^ Magnetization transfer imaging (MTI) detects signal indirectly from protons bound to macromolecules,^15^ which are not distinguishable on conventional MRI sequences. In healthy white matter (WM), this signal is dominated by myelin-bound protons, hence providing a degree of specificity for quantifying myelin content. The magnetization transfer ratio (*MTR*) has been extensively applied to the study of MS, although reported relationships between *MTR* and disability have varied.^16^ A recent meta-analysis which included RRMS cohorts of varying disease duration showed a negative association between NAWM *MTR* and Expanded Disability Status Scale (EDSS) scores,^16^ however many individual studies found no such relationship.^17–25^ The smaller number of analyses of NAGM *MTR* in mixed disease duration cohorts showed no significant association with EDSS.^16^ *MTR* data are confounded by T_1_ relaxation and B_1_^+^ magnetic field inhomogeneity effects, which may result in both increased variance and systematic bias in myelin estimation, and may partially account for inconsistent findings across different studies.

Magnetization transfer saturation (*MTsat*) signal incorporates an inherent T_1_ and B1+ correction,^26^ and is therefore thought to be more specific to myelin content compared with *MTR*. To date, *MTsat* has seen limited application in MS (Supplementary Table 1),^27–47^ although one cross-sectional study has reported associations between clinical disability and NAWM *MTsat*, but not *MTR*.^34^ The variable flip angle (VFA) T_1_ approximation required for *MTsat* calculation provides an additional quantitative measure of *T*_1_, which is also thought to be a proxy for myelin density.^48^ More marked focal *T*_1_ prolongation within WML provides a quantitative correlate of visible T_1_-weighted ‘black hole’ lesions, which are known to be associated with MS disease progression.^49^ The relationships between changes in myelin integrity in visible lesions and normal-appearing brain, measured using MTsat, disability progression, and interactions with radiological interval disease activity remain unexplored in early RRMS. Associations between *T*_1_ and minor disability worsening over one year were previously reported in the FutureMS cohort;^50^ disability measures over five years were not however previously available.

The primary aims of this study are to determine the degree to which demyelination, measured longitudinally *in vivo* using *MTsat* in NAWM and WML, (a) drives clinical disability progression in the five years following diagnosis of RRMS, (b) predicts future disability progression, and c) how the relationship between myelin integrity and disability relates to interval inflammatory disease activity reflected in NELs. As exploratory aims, we explore the association between early *T*_1_ changes and disability progression over five years in this early RRMS cohort, and re-examine previous analyses of disability evolution over one year using more stringent and widely-used definitions of disability worsening. We additionally examine associations between baseline myelin measures, diffusion-weighted imaging (dMRI)-derived metrics, blood-derived markers of neuroaxonal damage and disability progression, and we explore whether and how trajectories of five-year microstructural changes co-vary with disability progression.

## Materials and methods

### Participants

#### People with RRMS

Seventy-nine people with RRMS (pwRRMS, Figure 1) consented to take part in a single- centre (Anne Rowling Regenerative Neurology Centre, Edinburgh, United Kingdom) sub- study of FutureMS, a prospective multi-centre longitudinal inception cohort study of people recently diagnosed with RRMS in Scotland.^51,52^ For inclusion, participants had to be diagnosed with RRMS, according to 2010 McDonald criteria,^53^ up to six months prior to baseline visit, be over 18 years of age, and not yet be receiving treatment with any DMTs. Exclusion criteria included contraindication to MRI. Sample size for the wider FutureMS study was calculated based on simulation models,^54^ as previously described.^52^ The extended sub-study MRI protocol was pragmatically offered to all Edinburgh FutureMS participants once recruitment opened in November 2017 until recruitment ended across all centres in March 2019.^52^

**Figure 1:**
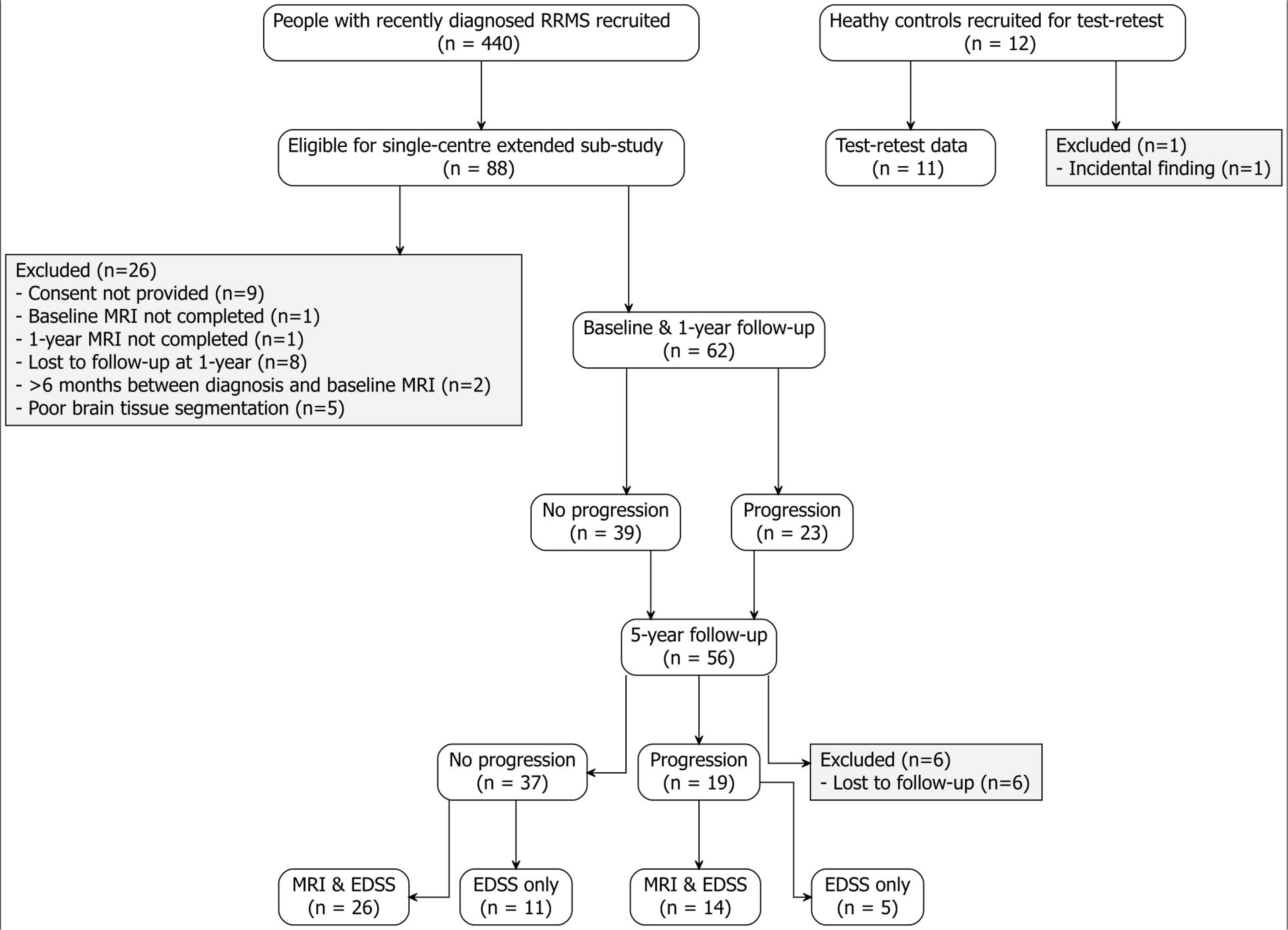
Participation flowchart for people with relapsing-remitting multiple sclerosis (RRMS) and healthy controls. Progression is defined relative to previous timepoint Expanded Disability Status Scale (EDSS) score (see main text for details).

#### Healthy controls

Twelve age- and sex-matched healthy controls were additionally recruited to establish test- retest agreement for each microstructural metric, as previously described.^37^ Exclusion criteria were contraindication to MRI and incidental findings on MRI. Control participants received an identical MRI protocol to pwRRMS, repeated within two weeks.

### Standard Protocol Approvals, Registrations, and Patient Consents

Ethics approval was obtained from the South East Scotland Research Ethics Committee 02 for the FutureMS sub-study (reference REC 15/SS/0233). The study conformed to the Declaration of Helsinki 2000 (amendments in 2002 and 2004) and Good Clinical Practice ICH guidelines. All participants provided written informed consent and data were anonymised before analysis.

### Clinical Assessment

Clinical assessment at baseline, one-year and five-year follow-up included EDSS score. The EDSS ranges from 0 to 10, with a higher score indicating more severe disability.^55^ Disability progression was defined as an increase of ≥1.5 points, ≥1.0 point or ≥0.5 points for EDSS scores of <1.0, 1.0 to 5.5, and ≥6.0, respectively, at the previous time point.

### MRI Acquisition

MRI acquisition took place at baseline, one-year, and five-year follow-up, as described previously.^37,52^ All imaging was obtained on a 3 Tesla Prisma MRI system (Siemens, Erlangen, Germany) with a 32-channel head coil. MRI data included: (1) a 3D T_1_-weighted Magnetization Prepared Rapid Acquisition Gradient Echo (MPRAGE) and (2) 2D dual echo (DE) for brain tissue segmentation; (3) 2D fluid-attenuated inversion recovery (FLAIR) for WML segmentation; (4) two 3D proton density fast low-angle shot spoiled gradient echo (SPGR) images with and without a Gaussian saturation pulse (9.984 ms; 500°) at 1.2 kHz offset from the water frequency, plus an additional 3D SPGR T_1_-weighted image required for *MTsat* and VFA *T*_1_ calculation; and, (4) multi-shell 2D diffusion-weighted spin-echo echo- planar imaging (151 directions; 0/200/500/1000/2000 s/mm2 b-values) with an additional three reverse phase-encoding volumes (b=0 s/mm2) to derive diffusion tensor imaging (DTI) and neurite orientation dispersion and density imaging (NODDI) parameters. Full MRI acquisition parameters are given in Supplementary Table 2.

### MRI Processing

#### Parametric maps

Following methodology detailed elsewhere,^37,50,52^ *MTsat* parametric maps (Figure 2) were calculated from MT_on_, MT_off_ and MT_T1w_ SPGR images (with echoes summed to increase signal-to-noise), and *T*_1_ was approximated from MT_off_ and MT_T1w_ images, according to equations described by Helms et al.^26^ in MATLAB (R2018b, software available: https://doi.org/10.7488/ds/2965).^59^

**Figure 2:**
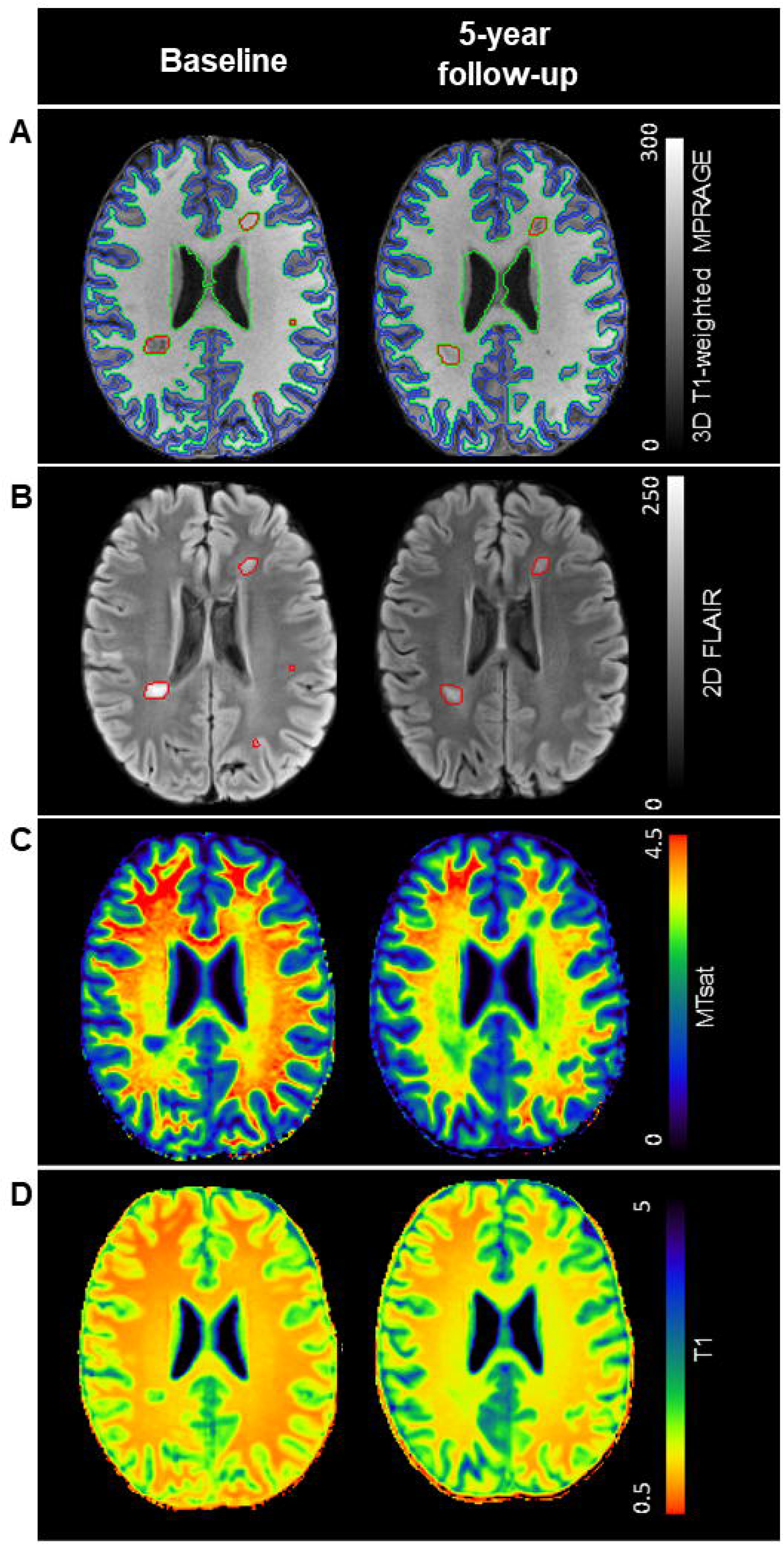
Example MR images for a FutureMS participant with recently diagnosed relapsing-remitting multiple sclerosis. Images show: (a) 3D T1-weighted MPRAGE with overlaid masks for normal-appearing white matter (green), grey matter (blue) and white matter lesions (WML, red); (b) 2D FLAIR with overlaid WML; (c) magnetization transfer saturation (MTsat), (d) and T1 parametric maps at study baseline and 5-year follow-up. Lower MTsat and higher T1 suggests reduced myelin.

DMRI data were processed with the NODDI toolbox^60^ (v1.0, mig.cs.ucl.ac.uk, MATLAB R2018b) and DTIFIT after performing brain extraction, removal of bulk motion and eddy- current induced distortions, and registration to first b0 volume with FSL (v6.0.1), as described previously.^52^

#### Brain tissue segmentation

As described in detail previously,^52^ skull-stripping was performed on DE proton density brain images with FSL^61^ (v6.0.1) and manually edited to obtain intracranial volumes (ICV). WML were segmented on T_2_ FLAIR (Figure 2), using an adapted thresholding method.^52,62^ Segmentation of brain tissue, including NAWM, cortical GM, deep GM (thalami, basal ganglia, and amygdala), and hippocampi, was performed on T_1_-weighted MPRAGE images with FreeSurfer (v6.0, https://surfer.nmr.mgh.harvard.edu/) with edited ICV masks and DE T_2_-weighted images as additional inputs. WML were subtracted from tissue masks and all other tissue masks were eroded by one voxel. Tissue volumes were calculated and corrected for ICV. GM and WM were similarly segmented for healthy control images.

To account for the influence of additional WML voxels forming over the follow-up period, baseline WML were registered to one- and five-year follow-up parametric maps using FSL flirt.

We also calculated the number of ‘prolonged *T*_1_ voxels’ in WML with *T*_1_ ≥2.00 s as detailed previously.^50^

Interval disease activity, evidenced by the presence of NELs at follow-up, was assessed by visual inspection on FLAIR by an experienced neuroradiologist.^52^

### Blood-derived markers

Blood plasma neurofilament light chain (NfL) and glial fibrillary acidic protein (GFAP) were measured using a multiplex single molecule array (Simoa) enzyme-linked immunosorbent assay (ELISA), as described previously.^35^ NfL and GFAP *z*-scores, adjusted for sex (GFAP only), age, and body mass index (BMI) were calculated using large reference populations.^56–58^

EBV nuclear antigen 1 (EBNA1) immunoglobulin G (IgG) titres were measured by an extended-dilution indirect ELISA using recombinant whole EBNA1 protein. Serum was assayed in duplicate across serial dilutions (1:200, 1:800, 1:3,200 and 1:12,800), with plate- specific normalisation using pooled control sera and on-plate controls. Quantitative titres were derived by fitting 4- or 5-parameter logistic dilution curves and calculating the reciprocal dilution corresponding to a predefined optical-density threshold; titres were log2- transformed for analysis.

### Statistical Analyses

Statistical analyses and extraction of regional median *MTsat* and *T*_1_ were carried out in RStudio (v1.4.1717, R v3.6.1).

#### Test-retest agreement

As a measure of test-retest agreement, and as previously described,^37^ Bland-Altman limits of agreements^63^ were established from healthy control WM and GM, between which 95% of repeated measurements would be expected to fall.

### Relationship with disability progression

#### Primary analyses

Primary analyses examined the association between change in regional *MTsat* between time points, and disability progression between the same and future time points was assessed with binomial logistic regression, adjusted for baseline age, sex, change in WML volume, baseline *MTsat*, and dichotomised DMT usage. The majority of participants were treated with low- efficacy DMTs or untreated at one-year (93.5%) and five-years (80.3%); there was thus limited scope to further interrogate the effect of high-efficacy treatment on associations.

False Discovery Rate (FDR) correction was performed. Overall goodness-of-fit was assessed with Nagelkerke’s^64^ (pseudo)-*R^2^* (*rms* package).^65^ Log-Likelihood Ratios with Wald’s chi- squared (*χ^2^*) tests and, when significant, receiver operating characteristic (ROC) curves with Akaike Information Criterion (AIC) were used to compare models against the null model and the reference model, i.e. with microstructural MRI markers removed.^66,67^ Odds ratios were calculated for coefficients of *z*-normalised variables. To determine the robustness of findings, analyses were repeated excluding the five observations with the largest Cook’s distance per model.

For significant primary models, to determine the influence of interval disease activity, models were repeated with inclusion of an interaction term between change over time in the given metric and NEL group.

#### Exploratory analyses

Analyses were repeated for *T*_1_, prolonged *T*_1_ voxel count in WML, NODDI, DTI, and baseline microstructural metrics to determine associations with disability progression. Baseline models included baseline WML volume, age and sex as covariates.

As NfL and GFAP are markers of neuroaxonal damage, and EBNA1 titres are elevated in people with MS, blood-derived markers were also added to primary analyses to determine incremental value.

To determine retrospectively whether the trajectory of microstructural changes differed between those who did or did not experience disability progression in the five years following diagnosis, participants with complete MRI and EDSS data were split into disability groups: ‘progressors’ who experienced disability progression between baseline and one-year, and between one-year and five-years; ‘early progressors’ who only showed progression between baseline and one-year; ‘late progressors’ who only progressed between one-year and five- years; and, ‘non-progressors’ with no evidence of progression between time points. Linear mixed-effects models, with baseline age, sex, baseline WML load, and DMT usage as covariates, were used to determine whether the microstructural metric significantly depends on the interaction term between progression group and time.

### Data availability

Researchers may request access to de-identified participant data from FutureMS. For information, please see www.annerowlingclinic.org/resource-access.

## Results

### Demographics

#### pwRRMS

Complete longitudinal *MTsat*, *T*_1_ and EDSS data were available for 62 pwRRMS at baseline and one-year, of whom 40 completed five-year MRI and EDSS (Figure 1 and Table 1), and 16 completed EDSS only (Supplementary Table 3).

The median change in EDSS for participants whose disability had progressed over one year was 1.0 point (range 0.5 to 3.5), and 1.5 points (range 0.5 to 3.0) for those who progressed from one-year to five-year follow-up.

In the 42 days prior to MRI, six pwRRMS at baseline, one pwRRMS at one-year follow-up, and one pwRRMS at five-years experienced relapses; only two pwRRMS at baseline received oral or intravenous steroid treatment in response to these relapses. The majority of participants (82.2%) experienced one or more clinical relapses in the baseline-to-one-year- follow-up interval period whereas only a minority of five-year participants (12.5%) experienced a relapse between one- and five-years.

Between baseline and one-year, 38 participants (61.2%) had initiated DMT, including dimethyl fumarate, (peg)interferon beta-1a, alemtuzumab, fingolimod, glatiramer acetate, and natalizumab. Between one- and five-year follow-up, 41 participants (73.2%) received at least one DMT, additionally including ocrelizumab, siponimod, cladribine, and diroximel fumarate.

#### Healthy controls

*MTsat* test-retest data was available for eleven healthy controls (seven female, mean [range] age 44 [27 to 58] years (Supplementary Table 4 and Figure 1 for exclusion reasons).

### Primary Analyses

#### *MTsat* associations with disability progression

##### Baseline-to-one-year disability progression

###### ‘Normal-appearing’ white matter

In NAWM, a greater decrease in *MTsat* between baseline and one-year was significantly associated with an increased likelihood of disability progression over the same time period (adjusted odds ratio [*aOR*] with *z*-normalised predictors [95% confidence interval] = 0.255 [0.078 to 0.602], *Wald χ^2^* = 7.15, FDR-corrected *p*-value = 0.015; Table 2; Figures 3A and 4A). Results did not materially change when the five most influential observations (determined by Cook’s distance) were removed (FDR-corrected *p*-value = 0.006). The model was more sensitive than the reference model with microstructural covariates removed (Area Under Curve [*AUC*] = 0.784 vs 0.645; Table 2 and Figure 4B). Despite significant groupwise effects, one-year change in NAWM *MTsat* for the majority of participants lay within test- retest limits of agreement (Supplementary Table 4-5 and Supplementary Results).

**Figure 3:**
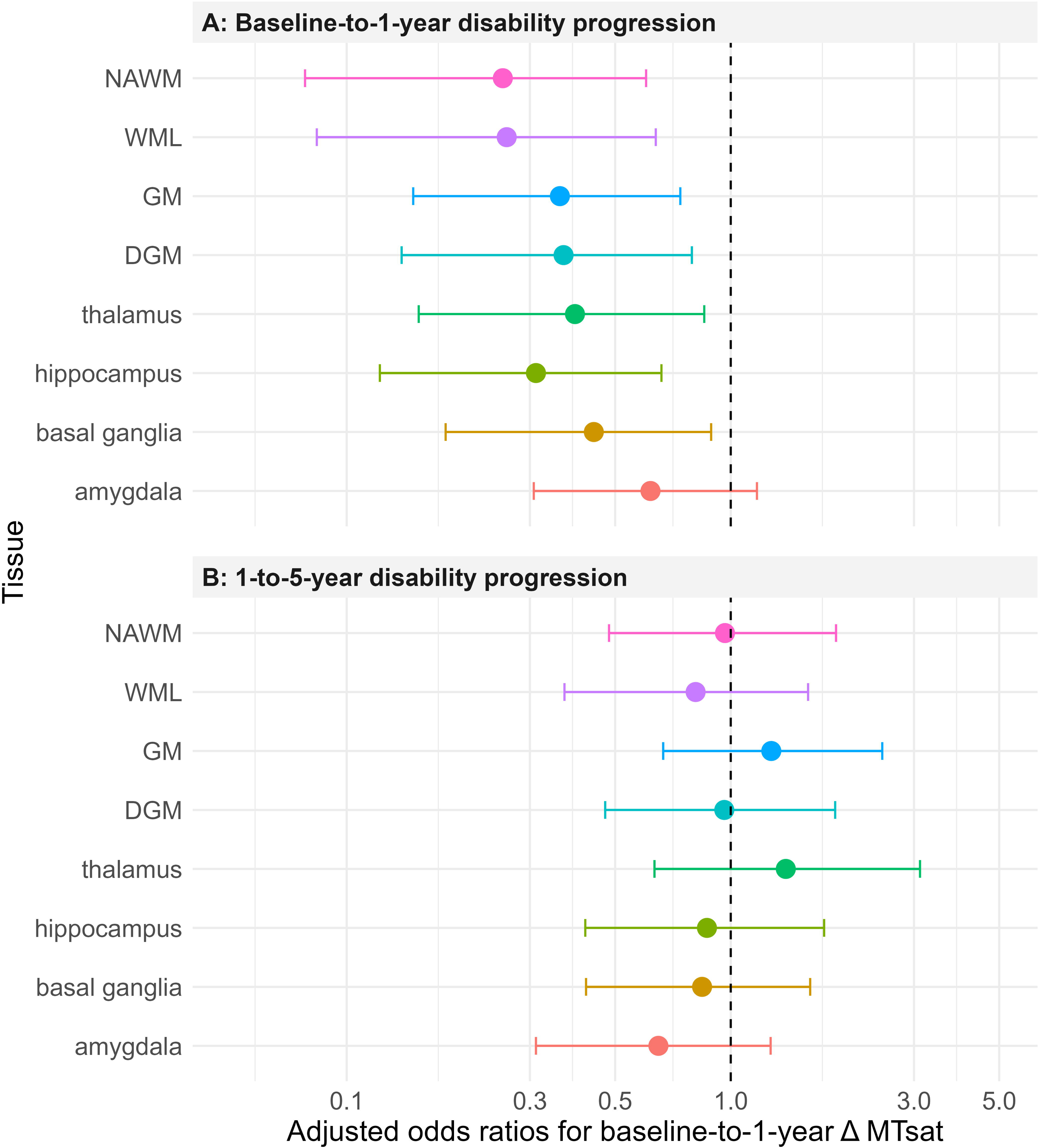
Forest plot showing adjusted odds ratios for baseline-to-one-year change in MTsat as a predictor of disability progression. Error bars indicate 95% confidence intervals for odds ratios (coloured points, log scale) of z-normalised one-year change in MTsat. Models were adjusted for age, sex, initiation of disease-modifying therapy and baseline MTsat. DGM: deep grey matter; GM: grey matter; MTsat: magnetization transfer saturation; NAWM: normal-appearing white matter; WML: white matter lesions present at baseline.

**Figure 4:**
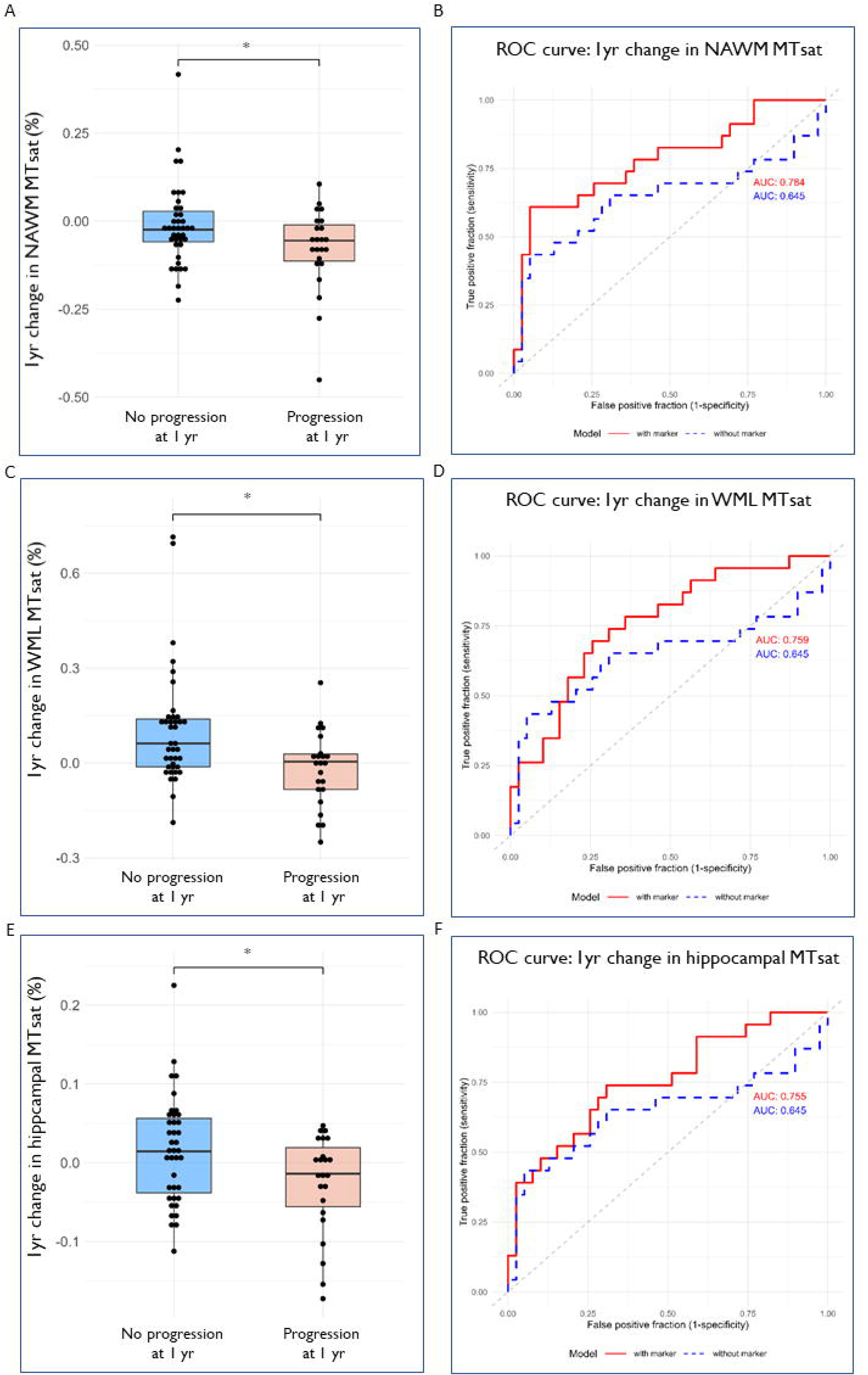
Relationship between one-year change in MTsat and disability progression over one-year in early relapsing-remitting MS. Boxplots show difference in (A) normal- appearing white matter (NAWM), (C) white matter lesions present at baseline (WML) and (E) hippocampal magnetization transfer saturation (MTsat, n = 62). Asterisks indicate significant difference between groups (FDR-corrected *P<0.05, covariate-adjusted binomial logistic regression). Corresponding receiver-operating characteristic (ROC) curves are shown for associations between one-year change in (B) NAWM, (D) WML and (F) hippocampal MTsat and one-year disability progression. Red lines represent model with all covariates; blue lines represent model with microstructural metrics removed. AUC: area under the curve.

###### White matter lesions

In WML, a greater decrease in *MTsat* over one year was significantly associated with disability progression (*aOR* = 0.261 [0.084 to 0.638], *χ^2^* = 6.79, FDR-corrected *p*-value = 0.015; Table 2; Supplementary Table 5; Figures 3A and 4C). Sensitivity analyses showed that this finding remained significant following exclusion of influential observations (FDR- corrected *p*-value = 0.006). The model with WML *MTsat* was more sensitive than the reference model (*AUC* = 0.759 vs 0.645; Table 2 and Figure 4D).

###### Other regions

Greater decreases in GM, deep GM, thalamic, basal ganglia, and hippocampal *MTsat* were significantly associated with one-year disability progression (FDR-corrected *p*-values < 0.05; Table 2 and Figure 3A), but changes in amygdala *MTsat* were not (FDR-corrected *p*-values = 0.151). Only the model with change in hippocampal *MTsat* outperformed both the null model and the reference model (Table 2 and Figure 4F). Sensitivity analyses however showed that, when the five most influential observations were removed, results remained significant and models outperformed the null and reference models.

##### One-to-five-year disability progression

One-to-five-year change in *MTsat* was not significantly associated with one-to-five-disability progression for any tissue region (FDR-corrected *p*-values >0.05; Supplementary Table 6).

### *MTsat* as a predictor of disability progression

#### One-to-five-year disability progression

Baseline-to-one-year change in *MTsat* did not predict change in disability progression between one-year and five-year follow-up (FDR-corrected *p*-values >0.05; Table 2; Figure 5).

**Figure 5:**
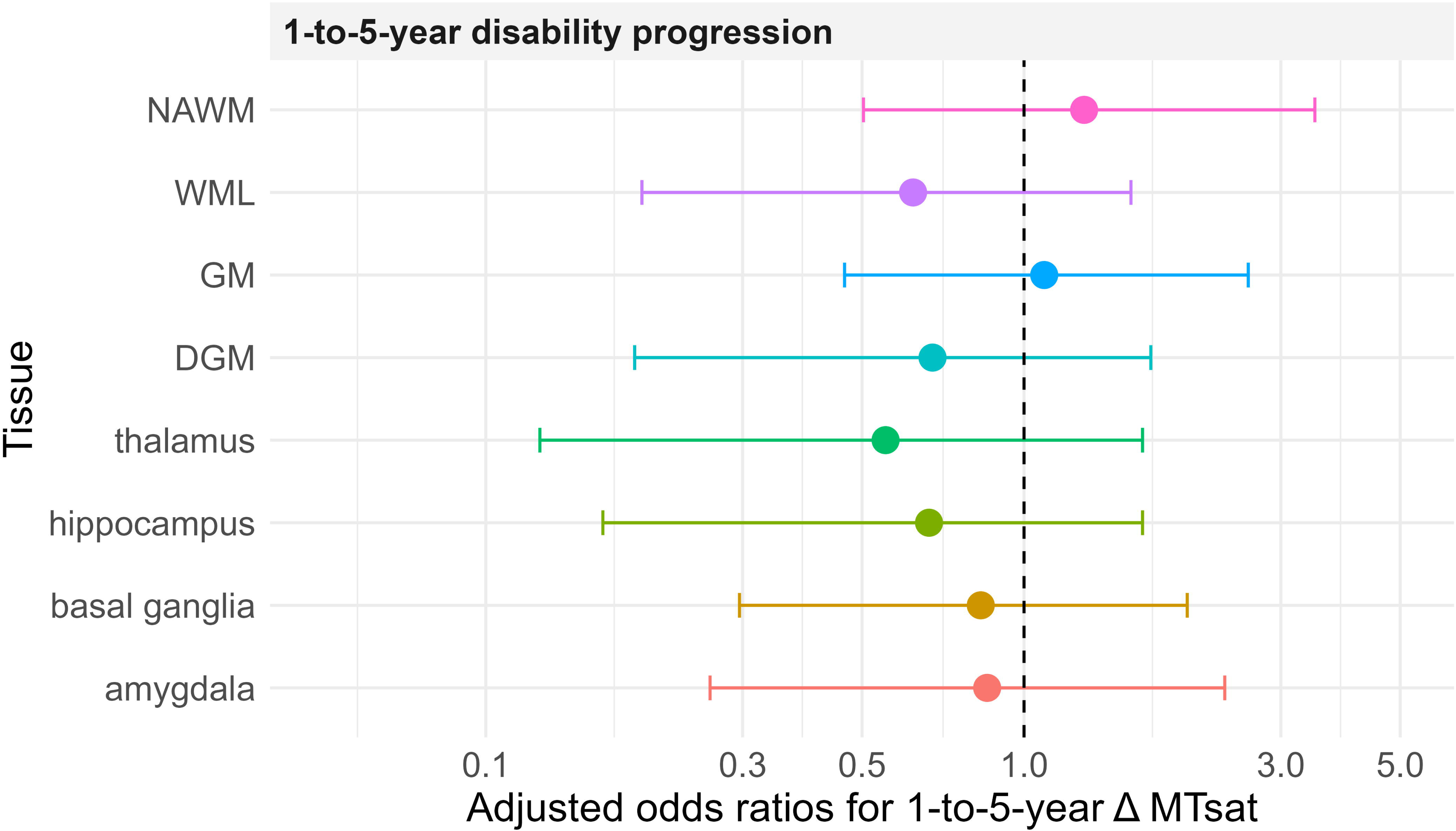
Forest plot showing adjusted odds ratios for one-to-five-year change in MTsat as a predictor of one-to-five-year disability progression. Error bars indicate 95% confidence intervals for odds ratios (coloured points, log scale) of *z*-normalised one-to-five- year change in MTsat. Models were adjusted for baseline age, sex, dichotomised disease- modifying therapy usage, and one-year MTsat. DGM: deep grey matter; GM: grey matter; MTsat: magnetization transfer saturation; NAWM: normal-appearing white matter; WML: white matter lesions present at baseline.

### Influence of NELs

*Post-hoc* analyses, adding the interaction term for change in *MTsat* and NEL grouping into significant models to determine the influence of interval radiological disease activity, showed no significant associations with disability progression for any tissue type (Supplementary Table 7).

### Exploratory Analyses

#### *T*_1_ associations with disability progression

##### Baseline-to-one-year disability progression

Neither baseline-to-one-year change in *T*_1_ nor change in the number of prolonged *T*_1_ WML voxels were significantly associated with baseline-to-one-year disability progression (FDR- corrected *p*-values >0.05; Supplementary Table 8).

##### One-to-five-year disability progression

Neither one-to-five-year change in *T*_1_ nor change in the number of prolonged *T*_1_ WML voxels were significantly associated with one-to-five-year disability progression (FDR-corrected *p*- values >0.05; Supplementary Table 9).

#### *T*_1_ as a predictor of disability progression

##### One-to-five-year disability progression

Baseline-to-one-year change in *T*_1_ did not significantly predict disability progression over the following four years (FDR-corrected *p*-values >0.05; Supplementary Table 8).

### Baseline predictors of clinical progression

Neither *MTsat* nor *T*_1_ at baseline was significantly associated with disability progression at five-year follow-up (Supplementary Table 10 and 11).

### DMRI associations with clinical progression

Neither baseline nor change in NODDI or DTI metrics were significantly associated with disability progression across the study period (Supplement Material 2).

### Blood-derived markers

Inclusion of NfL *z*-score, GFAP *z*-score, or EBNA-1 IgG titres in primary models for associations with disability progression did not improve model fit (Supplementary Material 3).

### Progression-associated differences in five-year microstructural trajectories

When retrospectively grouped according to progression trajectory over five years, there were four progressors (10%), nine early progressors (22.5%), ten late progressors (25%), and seventeen non-progressors (42.5%, Supplementary Table 12). Linear mixed-effects models showed that the interaction between progression grouping and time was significant for WML *MTsat* only, but this did not survive FDR correction (Table 3).

## Discussion

In this study, we investigate associations between quantitative microstructural MRI markers of myelin integrity and disability progression in the five years following RRMS diagnosis. We show that longitudinal groupwise one-year change in NAWM *MTsat* is associated with contemporaneous disability progression, but does not predict the subsequent four years of disability progression. We also demonstrate a relationship between decreasing *MTsat* in WML and disability progression over the first year, but not over subsequent one-to-five-year follow-up. We find no evidence to suggest that disability progression associated with early NAWM or WML demyelination is influenced by the presence of NELs, suggesting that changes in myelin integrity may occur independently of radiological inflammatory disease activity in recently diagnosed RRMS.

We also find evidence of progressive demyelination within GM and the majority of deep GM structures, which is detectable with *MTsat*, but generally show only modest associations with early disability progression.

Exploratory analyses show that the trajectory of WML *MTsat* nominally differs between individuals with different five-year disability progression trajectories; however, this finding does not survive FDR correction. Neither *T*_1_ nor any baseline metrics are associated with disability progression, and blood-derived NfL, GFAP and EBNA-1 markers did not provide incremental value to models.

### Longitudinal microstructural metrics and disability progression

#### Normal-appearing white matter

Diffuse NAWM abnormalities in pwMS, detected with quantitative MR techniques including *MTR* are well-documented;^68^ insights into the pathological drivers of disability divergence in early MS have, however, been limited by technique sensitivity and specificity. *MTsat* is thought to be more specific to myelin than *MTR* due to inherent correction for *T*_1_ relaxation effects (and, to some degree, *B*_1_^+^ inhomogeneities),^26^ and is strongly associated with myelin marker Luxol-Fast Blue.^69^ We previously observed a groupwise longitudinal decrease in *MTsat* in NAWM in our cohort of newly diagnosed pwRRMS, suggestive of widespread progressive demyelination.^37^ Here, we report for the first time that greater decreases in *MTsat* in NAWM in the year following RRMS diagnosis are associated with a significantly increased likelihood of early disability progression over the same one-year period. Our findings suggest that early subtle demyelination beyond visible WML contributes significantly to worsening disability.

The absence of any significant interaction between change in NAWM *MTsat* and NELs on disability evolution suggests that progressive ‘invisible’ myelin loss (demyelination and/or failure of remyelination) may be one of the substrates underlying progression in early RRMS, and occurs independently of MRI-visible inflammation. This may provide a potential mechanism underlying PIRA,^70^ and may have important treatment implications. The sample size of the current study, however, limits further robust interrogation of PIRA and relapse- associated worsening (RAW) phenotypes beyond the presence of NELs.

Our findings broadly support the hypothesis that disability in early MS is predominantly driven by a diffuse ‘smouldering’ pathological process, rather than by episodic focal inflammatory episodes.^71^ While paramagnetic rims around WML are thought to be a localised manifestation of chronic inflammation,^72^ widespread NAWM microglial activation, detected with PET and associated with microstructural disruption and disability, has also been reported in a mixed MS subtype cohort.^73^ It is not yet clear whether the widespread disability-relevant myelin loss demonstrated here results directly from more global brain inflammation in early RRMS.

Although one-year change in NAWM *MTsat* was associated with one-year disability progression, it was not predictive of one-to-five-year disability progression. One-to-five-year change in NAWM *MTsat* was also not associated with disability progression over that same period. There are a number of possible explanations for this. The rate of disability-relevant myelin loss may vary with time in MS. Indeed, we observed that the annualised *MTsat* change over the first year was more pronounced than over the subsequent four years. Trajectory over the first year following diagnosis is therefore not necessarily a predictor of disease behaviour over subsequent years. Variation in the efficacy of repair mechanisms in NAWM and treatment with DMTs during the one-to-five-year period may also mitigate longer term disability progression, and weaken any predictive relationship. Change in *MTsat* over five years is modest, and larger subject numbers or measurement over a longer period may be needed to identify statistically significant associations with and predictors of disability during this disease phase.

#### White matter lesions

Here, we demonstrate an association between changes in WML *MTsat* and disability progression over one year, although that change is not predictive of forward disability progression from one to five years. Within our recently diagnosed RRMS cohort, we previously noted a net increase in WML *MTsat* over time,^37^ which we speculated could be caused either by spontaneous remyelination after an acute inflammatory episode that prompted diagnosis, or reduction in oedema following acute inflammation in lesions; this accords with other evidence that remyelination within lesions may occur to a varying extent across patients.^74^ When grouped by EDSS progression trajectory however, we observe in this study that participants in our recently-diagnosed RRMS cohort with progression between time points tended to demonstrate a decrease in *MTsat* within WML, whilst an increase in WML *MTsat* was more commonly observed in those without progression. This suggests that recruitment of innate myelin repair mechanisms within focal WML may slow progression. Myelin-sensitive *MTR* within lesions has previously been applied as an endpoint in a remyelinating therapy trial,^75^ and *MTsat* may provide a more sensitive and specific end point for such studies in future.

The observation that *MTsat* changes in WML over a year do not predict one-to-five-year disability progression suggests that early spontaneous remyelination of already-extensively- damaged tissue does little to mitigate long-term disability accrual. The absence of detectable association with future disability progression could additionally be explained by technical *MTsat* ‘floor effects’ due to already very low myelin density within some WML.

#### Cortical grey matter

Previous studies have suggested that GM damage may contribute disproportionately to early disability progression in MS,^76–78^and cortical *MTR* changes indicative of myelin loss and repair measured over a year in a mixed MS cohort have recently been shown to predict disability after a five-year interval.^78^ In our early MS cohort, we found that cortical GM myelin changes measured with *MTsat* were significantly associated with one-year disability progression, which accords with previous findings. This model was, however, no better than the null (intercept-only) model, suggesting that GM may be marginally influential, but not the primary disability driver in the model. As with NAWM and WML, one-year cortical myelin change also did not predict one-to-five year disability progression and one-to-five year myelin changes were not associated with disability progression over the same period. Cortical GM is more sparsely myelinated than WM; low myelin density may reduce the accuracy of our myelin-sensitive metrics within the cortex. Poor detection of cortical GM lesions at 3 T clinical field strengths precludes reliable segmentation and therefore microstructural characterisation of focal cortical lesions. Our findings overall indicate that disability-relevant myelin changes occur within the cortex in early RRMS, but are difficult to detect without specific myelin-sensitized acquisition techniques, and may not predict longer term disability outcomes.

#### Deep grey matter

We observed a relationship between combined deep GM change in *MTsat* and one-year disability progression, in addition to individual associations with *MTsat* changes in the thalamus, basal ganglia and hippocampus. Histopathology has demonstrated demyelination in deep GM in MS.^79^ We are not aware of any study examining associations between longitudinal myelin changes within deep GM using *MTsat* and disability progression in recently-diagnosed RRMS. Our results illustrate that deep GM demyelination occurs at an early stage of disease and contributes to disability progression, albeit to a lesser extent than NAWM demyelination.

Although the thalamus is often classified as a GM structure, it is has complex internal anatomy and contains significant myelinated WM.^80^ A recent study showed no difference in two-year change in thalamic *MTsat*-derived myelin volume fraction (*MVF*) between healthy controls and pwRRMS of mixed disease duration.^42^ Previous work in early RRMS found that thalamic MTR was negatively associated with timepoint-averaged EDSS; the rate of *MTR* change was not associated with EDSS at two-year follow-up however.^81^ In later disease, reduced thalamic myelin density compared with healthy controls has been observed with *MTR*,^82,83^ and inhomogeneous (*ih*)*MTR*.^83^ Our finding of a relationship between thalamic *MTsat* and disability progression over one year, but not five years, broadly accords with these findings. Although myelin loss within the thalamus appears relevant to early disability progression, our results do not support the view that early thalamic demyelination predicts sustained disability progression over a longer time period.

In the basal ganglia, which comprise the caudate, putamen and pallidum, greater demyelination was associated with one-year disability progression in our early RRMS cohort. We are not aware of any other studies examining disability progression and demyelination measured with *MTsat* within the basal ganglia, although some evidence supports cross- sectional associations between disability and myelin proton fraction (*MPF*) within the caudate nucleus and putamen in MS.^84^ Basal ganglia demyelination however did not differentiate between those who did and did not progress at five-year follow-up, and is therefore not a stratifier of future disease trajectory.

Decreasing *MTsat* in the hippocampus appeared to significantly increase the likelihood of disability progression at one year in our newly-diagnosed RRMS cohort. Hippocampal demyelination is a known histopathological feature of MS,^85^ and significant associations between EDSS and myelin-sensitive *χ*-separation quantitative susceptibility dependent MRI of the hippocampi have been reported.^86^ Lower hippocampal sub-field volumes have also been reported in early RRMS compared with healthy controls^87^ and with MS diagnosis in people with clinically isolated syndrome (CIS).^88^ There are, however, a number of confounds that may contribute to our findings. The morphology of the hippocampus and adjacent CSF and choroid plexus in the temporal horn of the lateral ventricle make reliable segmentation difficult, and a small number of contributing voxels and partial volume effects from adjacent structures make *MTsat* signal vulnerable to subtle volume change. Moreover, susceptibility effects from the sphenoid bone and sinus may also influence signal return. The hippocampus primarily subserves memory function, and mechanisms underlying any direct causal relationship between hippocampal dysfunction and EDSS which is substantially motor- weighted should also be considered. Cognitive testing of specific memory domains is needed to elucidate any relationship between early hippocampal demyelination and progression of memory deficits, which appear more prevalent in MS with longer disease duration.^89^ Although our findings suggest possible disability-relevant hippocampal changes in RRMS, we therefore interpret these with caution.

### Baseline microstructural metrics and disability progression

We found no significant association between baseline *MTsat* and subsequent disability progression. This accords with previous cross-sectional studies which found no association between myelin measures and EDSS;^28,30,32^ although those studies examined EDSS at a single time point rather than change over time.

Our data from a sizeable cohort of recently diagnosed pwRRMS support the view that a single time point MRI measure of microstructural integrity is insufficient to predict EDSS clinical disability evolution over one year or five years. It is, however, possible that such associations may become apparent with longer follow-up, or with the development of more nuanced disability measures.

### *T*_1_ measures

In contrast to *MTsat*, we found no evidence of associations between disability and NAWM *T*_1,_ nor of prolonged *T*_1_ WML voxels and disability progression in our exploratory analyses. While we have previously reported a relationship between NAWM *T*_1_ and prolonged *T*_1_ WML and EDSS disability progression in our cohort using a ≥ 0.5 point threshold,^50^ here we show that, unlike associations with *MTsat*, this relationship does not persist with more stringent clinical criteria. We also previously reported relationships between longitudinal *T*_1_ measures in deep GM and one-year disability progression;^50^ again, these are not seen with more stringent progression criteria or as predictors of five-year disability worsening.

Floor effects have been observed previously with *T*_1_ prolongation in lesions,^48^ which may contribute to our null findings, and *T*_1_ changes in sparsely-myelinated cortex are likely to be dominated by factors other than myelin density.

We have previously reported longitudinal diffusion tensor imaging (DTI) or Neurite Orientation Dispersion and Density Imaging^60^ findings over one year in this cohort,^37^ and note that we also found no significant relationships between disability progression and any diffusion metrics for any brain region (Supplementary Material 2).

Our current results suggest that *MTsat* is a more sensitive measure than *T*_1_ for detecting subtle diffuse demyelination that causes more pronounced and longer-term disability progression. *MTsat* may therefore be more appropriate as a clinical trial surrogate endpoint, using widely accepted definitions of disability progression.

### Limitations

The FutureMS study was designed to recruit a cohort at a homogeneous disease stage; variation in recruitment with regard to the time of first reported symptom onset was not explicitly included in our analyses due to the poor reliability of determining first symptom onset compared with date of diagnosis, and a strong association with age. We also did not account for possible bias introduced by exclusion of subjects at recruitment, nor due to missing data; it is difficult to establish in which direction this may have biased our conclusions.

We acknowledge the limitations of the EDSS as a measure of disability, including the non- continuous and non-normal distribution of scores within MS cohorts.^67^ More nuanced, longitudinal cognitive testing, particularly with respect to memory deficits, may be required to fully elucidate the contribution of deep GM demyelination to disability progression.

All participants were treatment-naïve at baseline, and dichotomised use of DMTs between time points was included as a covariate in our models. FutureMS is, however, a ‘real world’ observational cohort and participants may initiate or switch DMTs between time points for a variety of reasons (including interval disease activity, risk profiles and adverse effects).

Explicit *B*_1_^+^ correction was not performed; MTsat, however, inherently corrects *B*_1_^+^ inhomogeneities to some degree. Although longitudinal measurements will mitigate the impact of residual *B*_1_^+^ inhomogeneities, and the occurrence of a systematic difference in *B*_1_^+^ distribution between disability groups is unlikely, the accuracy of cross-sectional *T*_1_, and to a certain extent *MTsat*, may be reduced. Our future studies will include acquisition of *B*_1_^+^ maps.

Lesion filling prior to brain tissue segmentation was not performed. Comparisons between segmentations performed with and without lesion filling (data not shown) demonstrate the impact on resulting brain tissue masks is minimal-to-none.

Understanding technique reproducibility is important for establishing sensitivity to longitudinal biological change. While changes in *MTsat* over one year were significant at the group level, test-retest limits of agreement established in healthy control WM indicate that they will not necessarily distinguish between individuals with stable/improving and those with worsening disability (see Supplementary Results). Moreover, technique reproducibility measured in healthy control data over a two-week period may underestimate test-retest variability over the one-year interval studied here in pwRRMS.

## Conclusion

Ongoing diffuse NAWM demyelination detected by longitudinal myelin-sensitive *MTsat* imaging is associated with one-year disability progression in early RRMS, independently of interval radiological disease activity. Widespread ongoing disability-relevant demyelination in WML and GM is also seen over the year following MS diagnosis. None of our microstructural measures predict future disability progression, and indeed myelin changes over the subsequent four years were modest and not associated with disability progression. Our findings provide potential mechanisms driving early disability progression beyond NELs, and suggest that microstructural evaluation of NAWM (and perhaps GM) may be at least as important as WML features for assessing disease trajectory. Quantitative myelin measures therefore provide an important adjunct to conventional indicators of activity for assessing otherwise ‘hidden’ disease evolution and therapeutic response in RRMS. Greater myelin changes over the first year but not subsequently suggest a disease tempo change and point towards a potentially important window of therapeutic opportunity. *MTsat* shows promise as a clinically-applicable disease stratifier and surrogate endpoint in future therapeutic trials, especially those targeted at preventing progression in those without radiological inflammatory disease activity.

## Supporting information

STROBE

## Acknowledgements

With special thanks to all participants in FutureMS, the radiographers at the Royal Infirmary of Edinburgh, Dr Daisy Mollison, and Edinburgh Imaging. We would also like to thank other non-author contributors of the FutureMS consortium listed here: https://edin.ac/4ePeDl3.

## Abbreviations

AIC: Akaike information criterion
aOR: Adjusted odds ratio
CSF: Cerebrospinal fluid
DMTs: Disease-modifying therapies
DE: Dual echo
EDSS: Expanded Disability Status Scale
FDR: False discovery rate
FLAIR: Fluid-attenuated inversion recovery
GM: Grey matter
ICV: Intracranial volume
MPRAGE: Magnetization Prepared Rapid Acquisition Gradient Echo
MS: Multiple sclerosis
MTI: Magnetization transfer imaging
MTR: Magnetization transfer ratio
MTsat: Magnetization transfer saturation
NAWM: ‘Normal-appearing’ white matter
NEL: New or enlarging white matter lesions
PIRA: Progression independent of relapse activity
ROC: Receiver operating characteristic
(pw)RRMS: (People with) relapsing-remitting multiple sclerosis
SPGR: Spoiled gradient echo
WM: White matter
WML: White matter lesions

## References

1. Trapp BD, Peterson J, Ransohoff RM, Rudick R, Mork S, Bo L. Axonal transection in the lesions of multiple sclerosis. N Engl J Med. Jan 29 1998;338(5):278–85. doi:10.1056/NEJM199801293380502

2. Absinta M, Lassmann H, Trapp BD. Mechanisms underlying progression in multiple sclerosis. Curr Opin Neurol. Jun 2020;33(3):277–285. doi:10.1097/WCO.0000000000000818

3. Wiendl H, Gold R, Berger T, et al. Multiple Sclerosis Therapy Consensus Group (MSTCG): position statement on disease-modifying therapies for multiple sclerosis (white paper). Ther Adv Neurol Disord. 2021;14:17562864211039648. doi:10.1177/17562864211039648

4. Muller J, Cagol A, Lorscheider J, et al. Harmonizing Definitions for Progression Independent of Relapse Activity in Multiple Sclerosis: A Systematic Review. JAMA Neurol. Nov 1 2023;80(11):1232–1245. doi:10.1001/jamaneurol.2023.3331

5. Thompson AJ, Banwell BL, Barkhof F, et al. Diagnosis of multiple sclerosis: 2017 revisions of the McDonald criteria. Lancet Neurol. Feb 2018;17(2):162–173. doi:10.1016/S1474-4422(17)30470-2

6. Barkhof F. MRI in multiple sclerosis: correlation with expanded disability status scale (EDSS). Mult Scler. Aug 1999;5(4):283–6. doi:10.1177/135245859900500415

7. Chard D, Trip SA. Resolving the clinico-radiological paradox in multiple sclerosis. F1000Res. 2017;6:1828. doi:10.12688/f1000research.11932.1

8. Magliozzi R, Reynolds R, Calabrese M. MRI of cortical lesions and its use in studying their role in MS pathogenesis and disease course. Brain Pathol. Sep 2018;28(5):735–742. doi:10.1111/bpa.12642

9. Sastre-Garriga J, Pareto D, Battaglini M, et al. MAGNIMS consensus recommendations on the use of brain and spinal cord atrophy measures in clinical practice. Nat Rev Neurol. Mar 2020;16(3):171–182. doi:10.1038/s41582-020-0314-x

10. Meijboom R, York EN, Kampaite A, et al. Patterns of brain atrophy in recently- diagnosed relapsing-remitting multiple sclerosis. PLoS One. 2023;18(7):e0288967. doi:10.1371/journal.pone.0288967

11. Tsivgoulis G, Katsanos AH, Grigoriadis N, et al. The effect of disease modifying therapies on brain atrophy in patients with relapsing-remitting multiple sclerosis: a systematic review and meta-analysis. PLoS One. 2015;10(3):e0116511. doi:10.1371/journal.pone.0116511

12. Cagol A, Schaedelin S, Barakovic M, et al. Association of Brain Atrophy With Disease Progression Independent of Relapse Activity in Patients With Relapsing Multiple Sclerosis. JAMA Neurol. Jul 1 2022;79(7):682–692. doi:10.1001/jamaneurol.2022.1025

13. Davies GR, Hadjiprocopis A, Altmann DR, et al. Normal-appearing grey and white matter T1 abnormality in early relapsing-remitting multiple sclerosis: a longitudinal study. Mult Scler. Mar 2007;13(2):169–77. doi:10.1177/1352458506070726

14. Granziera C, Wuerfel J, Barkhof F, et al. Quantitative magnetic resonance imaging towards clinical application in multiple sclerosis. Brain. Jun 22 2021;144(5):1296–1311. doi:10.1093/brain/awab029

15. Wolff SD, Balaban RS. Magnetization transfer contrast (MTC) and tissue water proton relaxation in vivo. Magn Reson Med. Apr 1989;10(1):135–44. doi:10.1002/mrm.1910100113

16. York EN, Thrippleton MJ, Meijboom R, Hunt DPJ, Waldman AD. Quantitative magnetization transfer imaging in relapsing-remitting multiple sclerosis: a systematic review and meta-analysis. Brain Commun. 2022;4(2):fcac088. doi:10.1093/braincomms/fcac088

17. Amann M, Sprenger T, Naegelin Y, et al. Comparison between balanced steady-state free precession and standard spoiled gradient echo magnetization transfer ratio imaging in multiple sclerosis: methodical and clinical considerations. Neuroimage. Mar 2015;108:87–94. doi:10.1016/j.neuroimage.2014.12.045

18. Bellmann-Strobl J, Stiepani H, Wuerfel J, et al. MR spectroscopy (MRS) and magnetisation transfer imaging (MTI), lesion load and clinical scores in early relapsing remitting multiple sclerosis: a combined cross-sectional and longitudinal study. European radiology. Aug 2009;19(8):2066–74. doi:10.1007/s00330-009-1364-z

19. Catalaa I, Grossman RI, Kolson DL, et al. Multiple sclerosis: magnetization transfer histogram analysis of segmented normal-appearing white matter. Radiology. Aug 2000;216(2):351–5. doi:10.1148/radiology.216.2.r00au16351

20. Davies GR, Ramio-Torrenta L, Hadjiprocopis A, et al. Evidence for grey matter MTR abnormality in minimally disabled patients with early relapsing-remitting multiple sclerosis. J Neurol Neurosurg Psychiatry. Jul 2004;75(7):998–1002. doi:10.1136/jnnp.2003.021915

21. Davies GR, Altmann DR, Hadjiprocopis A, et al. Increasing normal-appearing grey and white matter magnetisation transfer ratio abnormality in early relapsing-remitting multiple sclerosis. J Neurol. Sep 2005;252(9):1037–44. doi:10.1007/s00415-005-0808-x

22. Mangia S, Carpenter AF, Tyan AE, Eberly LE, Garwood M, Michaeli S. Magnetization transfer and adiabatic T1rho MRI reveal abnormalities in normal-appearing white matter of subjects with multiple sclerosis. Mult Scler. Jul 2014;20(8):1066–73. doi:10.1177/1352458513515084

23. Siemonsen S, Young KL, Bester M, et al. Chronic T2 Lesions in Multiple Sclerosis are Heterogeneous Regarding Phase MR Imaging. Clinical neuroradiology. Dec 2016;26(4):457–464. doi:10.1007/s00062-015-0389-8

24. Zhang L, Wen B, Chen T, et al. A comparison study of inhomogeneous magnetization transfer (ihMT) and magnetization transfer (MT) in multiple sclerosis based on whole brain acquisition at 3.0 T. Magnetic resonance imaging. Jul 2020;70:43–49. doi:10.1016/j.mri.2020.03.010

25. Zivadinov R, Hussein S, Stosic M, et al. Glatiramer acetate recovers microscopic tissue damage in patients with multiple sclerosis. A case-control diffusion imaging study. Pathophysiology. Feb 2011;18(1):61–8. doi:10.1016/j.pathophys.2010.04.007

26. Helms G, Dathe H, Kallenberg K, Dechent P. High-resolution maps of magnetization transfer with inherent correction for RF inhomogeneity and T1 relaxation obtained from 3D FLASH MRI. Magn Reson Med. Dec 2008;60(6):1396–407. doi:10.1002/mrm.21732

27. Berg RC, Menegaux A, Amthor T, et al. Comparing myelin-sensitive magnetic resonance imaging measures and resulting g-ratios in healthy and multiple sclerosis brains. Neuroimage. Dec 1 2022;264:119750. doi:10.1016/j.neuroimage.2022.119750

28. Elkady AM, Wu Z, Leppert IR, Arnold DL, Narayanan S, Rudko DA. Assessing the differential sensitivities of wave-CAIPI ViSTa myelin water fraction and magnetization transfer saturation for efficiently quantifying tissue damage in MS. Mult Scler Relat Disord. Nov 2021;56:103309. doi:10.1016/j.msard.2021.103309

29. Lommers E, Guillemin C, Reuter G, et al. Voxel-Based quantitative MRI reveals spatial patterns of grey matter alteration in multiple sclerosis. Hum Brain Mapp. Mar 2021;42(4):1003–1012. doi:10.1002/hbm.25274

30. Lommers E, Simon J, Reuter G, et al. Multiparameter MRI quantification of microstructural tissue alterations in multiple sclerosis. Neuroimage Clin. 2019;23:101879. doi:10.1016/j.nicl.2019.101879

31. Campbell JSW, Leppert IR, Narayanan S, et al. Promise and pitfalls of g-ratio estimation with MRI. Neuroimage. Nov 15 2018;182:80–96. doi:10.1016/j.neuroimage.2017.08.038

32. Saccenti L, Hagiwara A, Andica C, et al. Myelin Measurement Using Quantitative Magnetic Resonance Imaging: A Correlation Study Comparing Various Imaging Techniques in Patients with Multiple Sclerosis. Cells. Feb 8 2020;9(2)doi:10.3390/cells9020393

33. Tagge IJ, Leppert IR, Fetco D, et al. Permanent tissue damage in multiple sclerosis lesions is associated with reduced pre-lesion myelin and axon volume fractions. Mult Scler. Nov 2022;28(13):2027–2037. doi:10.1177/13524585221110585

34. Lema A, Bishop C, Malik O, et al. A Comparison of Magnetization Transfer Methods to Assess Brain and Cervical Cord Microstructure in Multiple Sclerosis. J Neuroimaging. Mar 2017;27(2):221–226. doi:10.1111/jon.12377

35. York EN, Martin SJ, Meijboom R, et al. MRI-derived g-ratio and lesion severity in newly diagnosed multiple sclerosis. Brain Commun. 2021;3(4):fcab249. doi:10.1093/braincomms/fcab249

36. Rahmanzadeh R, Weigel M, Lu PJ, et al. A comparative assessment of myelin- sensitive measures in multiple sclerosis patients and healthy subjects. Neuroimage Clin. Aug 30 2022;36:103177. doi:10.1016/j.nicl.2022.103177

37. York EN, Meijboom R, Thrippleton MJ, et al. Longitudinal microstructural MRI markers of demyelination and neurodegeneration in early relapsing-remitting multiple sclerosis: Magnetisation transfer, water diffusion and g-ratio. Neuroimage Clin. 2022;36:103228. doi:10.1016/j.nicl.2022.103228

38. Longoni G, Martinez Chavez E, Young K, et al. Magnetization transfer saturation reveals subclinical optic nerve injury in pediatric-onset multiple sclerosis. Mult Scler. Feb 2023;29(2):212–220. doi:10.1177/13524585221137500

39. Vandeleene N, Guillemin C, Dauby S, et al. Using quantitative magnetic resonance imaging to track cerebral alterations in multiple sclerosis brain: A longitudinal study. Brain Behav. May 2023;13(5):e2923. doi:10.1002/brb3.2923

40. Guillemin C, Vandeleene N, Charonitis M, et al. Brain microstructure is linked to cognitive fatigue in early multiple sclerosis. J Neurol. Mar 28 2024;doi:10.1007/s00415-024-12316-1

41. Sanabria-Diaz G, Cagol A, Lu PJ, et al. Advanced MRI Measures of Myelin and Axon Volume Identify Repair in Multiple Sclerosis. Ann Neurol. Oct 10 2024;doi:10.1002/ana.27102

42. Cagol A, Ocampo-Pineda M, Lu PJ, et al. Advanced Quantitative MRI Unveils Microstructural Thalamic Changes Reflecting Disease Progression in Multiple Sclerosis. Neurology(R) neuroimmunology & neuroinflammation. Nov 2024;11(6):e200299. doi:10.1212/NXI.0000000000200299

43. Mastantuono M, Cagol A, Ocampo-Pineda M, et al. Choroid plexus morphological and microstructural changes across inflammatory phenotypes in multiple sclerosis. Mult Scler. Jul 21 2026:13524585261462513. doi:10.1177/13524585261462513

44. Wright SJ, McKay SG, Bou JR, et al. T(1) Over Squared Proton Density Ratio to Characterize Multiple Sclerosis Lesions. Ann Clin Transl Neurol. Jun 7 2026;doi:10.1002/acn3.70446

45. Schlaeger S, Muhlau M, Gilbert G, et al. Sensitivity of multi-parametric quantitative magnetic resonance imaging for multiple sclerosis pathology. PLoS One. 2025;20(4):e0318415. doi:10.1371/journal.pone.0318415

46. Thevakumaran R, Blinder S, Couch M, et al. Subcortical microglial inflammation is uniquely linked to subpial cortical demyelination in multiple sclerosis. Neuroimage Clin. 2026;49:103962. doi:10.1016/j.nicl.2026.103962

47. Trang H, Hartung TJ, Chen Q, et al. A quantitative multi-parameter mapping protocol standardized for clinical research in multiple sclerosis. Sci Rep. Dec 16 2024;14(1):30481. doi:10.1038/s41598-024-80274-9

48. Kolb H, Absinta M, Beck ES, et al. 7T MRI Differentiates Remyelinated from Demyelinated Multiple Sclerosis Lesions. Ann Neurol. Oct 2021;90(4):612–626. doi:10.1002/ana.26194

49. Truyen L, van Waesberghe JH, van Walderveen MA, et al. Accumulation of hypointense lesions (“black holes”) on T1 spin-echo MRI correlates with disease progression in multiple sclerosis. Neurology. Dec 1996;47(6):1469–76. doi:10.1212/wnl.47.6.1469

50. Harper JG, York EN, Meijboom R, et al. Quantitative T(1) brain mapping in early relapsing-remitting multiple sclerosis: longitudinal changes, lesion heterogeneity and disability. European radiology. Jun 2024;34(6):3826–3839. doi:10.1007/s00330-023-10351-6

51. Kearns PKA, Martin SJ, Chang J, et al. FutureMS cohort profile: a Scottish multicentre inception cohort study of relapsing-remitting multiple sclerosis. BMJ Open. Jun 29 2022;12(6):e058506. doi:10.1136/bmjopen-2021-058506

52. Meijboom R, Wiseman SJ, York EN, et al. Rationale and design of the brain magnetic resonance imaging protocol for FutureMS: a longitudinal multi-centre study of newly diagnosed patients with relapsing-remitting multiple sclerosis in Scotland. Wellcome Open Res. 2022;7:94. doi:10.12688/wellcomeopenres.17731.1

53. Polman CH, Reingold SC, Banwell B, et al. Diagnostic criteria for multiple sclerosis: 2010 revisions to the McDonald criteria. Ann Neurol. Feb 2011;69(2):292–302. doi:10.1002/ana.22366

54. Dobbin KK, Simon RM. Sample size planning for developing classifiers using high- dimensional DNA microarray data. Biostatistics. Jan 2007;8(1):101–17. doi:10.1093/biostatistics/kxj036

55. Kurtzke JF. Rating neurologic impairment in multiple sclerosis: an expanded disability status scale (EDSS). Neurology. Nov 1983;33(11):1444–52. doi:10.1212/wnl.33.11.1444

56. Benkert P, Meier S, Schaedelin S, et al. Serum neurofilament light chain for individual prognostication of disease activity in people with multiple sclerosis: a retrospective modelling and validation study. Lancet Neurol. Mar 2022;21(3):246–257. doi:10.1016/S1474-4422(22)00009-6

57. Meier S, Willemse EAJ, Schaedelin S, et al. Serum Glial Fibrillary Acidic Protein Compared With Neurofilament Light Chain as a Biomarker for Disease Progression in Multiple Sclerosis. JAMA Neurol. Mar 1 2023;80(3):287–297. doi:10.1001/jamaneurol.2022.5250

58. Benkert P, Maleska Maceski A, Schaedelin S, et al. Serum Glial Fibrillary Acidic Protein and Neurofilament Light Chain Levels Reflect Different Mechanisms of Disease Progression under B-Cell Depleting Treatment in Multiple Sclerosis. Ann Neurol. Oct 16 2024;97(1):104–15. doi:10.1002/ana.27096

59. Magnetisation transfer saturation (MTsat) processing. 10.7488/ds/2965. Software: University of Edinburgh. Centre for Clinical Brain Sciences. doi: 10.7488/ds/2965

60. Zhang H, Schneider T, Wheeler-Kingshott CA, Alexander DC. NODDI: practical in vivo neurite orientation dispersion and density imaging of the human brain. Neuroimage. Jul 16 2012;61(4):1000–16. doi:10.1016/j.neuroimage.2012.03.072

61. Jenkinson M, Beckmann CF, Behrens TE, Woolrich MW, Smith SM. Fsl. Neuroimage. Aug 15 2012;62(2):782–90. doi:10.1016/j.neuroimage.2011.09.015

62. Zhan TM, Zhan YZ, Liu Z, Xiao L, Wei ZH. Automatic method for white matter lesion segmentation based on T1-fluid-attenuated inversion recovery images. Iet Computer Vision. Aug 2015;9(4):447–455. doi:10.1049/iet-cvi.2014.0121

63. Bland JM, Altman DG. Statistical Methods for Assessing Agreement between Two Methods of Clinical Measurement. Lancet. Feb 8 1986;1(8476):307–310. doi:DOI 10.1016/s0140-6736(86)90837-8

64. Nagelkerke NJD. A Note on a General Definition of the Coefficient of Determination. Biometrika. Sep 1991;78(3):691–692. doi:DOI 10.1093/biomet/78.3.691

65. Harrell Jr FE. rms: Regression modeling strategies. R package version. 2016;5(2)

66. Kunchok A, Lechner-Scott J, Granella F, et al. Prediction of on-treatment disability worsening in RRMS with the MAGNIMS score. Mult Scler. Apr 2021;27(5):695–705. doi:10.1177/1352458520936823

67. van Munster CE, Uitdehaag BM. Outcome Measures in Clinical Trials for Multiple Sclerosis. CNS Drugs. Mar 2017;31(3):217–236. doi:10.1007/s40263-017-0412-5

68. Miller DH, Thompson AJ, Filippi M. Magnetic resonance studies of abnormalities in the normal appearing white matter and grey matter in multiple sclerosis. J Neurol. Dec 2003;250(12):1407–19. doi:10.1007/s00415-003-0243-9

69. Georgiadis M, Schroeter A, Gao Z, et al. Nanostructure-specific X-ray tomography reveals myelin levels, integrity and axon orientations in mouse and human nervous tissue. Nat Commun. May 19 2021;12(1):2941. doi:10.1038/s41467-021-22719-7

70. Lublin FD, Haring DA, Ganjgahi H, et al. How patients with multiple sclerosis acquire disability. Brain. Sep 14 2022;145(9):3147–3161. doi:10.1093/brain/awac016

71. Giovannoni G, Popescu V, Wuerfel J, et al. Smouldering multiple sclerosis: the ‘real MS’. Ther Adv Neurol Disord. 2022;15:17562864211066751. doi:10.1177/17562864211066751

72. Dal-Bianco A, Grabner G, Kronnerwetter C, et al. Slow expansion of multiple sclerosis iron rim lesions: pathology and 7 T magnetic resonance imaging. Acta Neuropathol. Jan 2017;133(1):25–42. doi:10.1007/s00401-016-1636-z

73. Bezukladova S, Tuisku J, Matilainen M, et al. Insights into disseminated MS brain pathology with multimodal diffusion tensor and PET imaging. Neurology(R) neuroimmunology & neuroinflammation. May 2020;7(3)doi:10.1212/NXI.0000000000000691

74. Patrikios P, Stadelmann C, Kutzelnigg A, et al. Remyelination is extensive in a subset of multiple sclerosis patients. Brain. Dec 2006;129(Pt 12):3165–72. doi:10.1093/brain/awl217

75. Brown JWL, Prados F, Altmann DR, et al. Remyelination varies between and within lesions in multiple sclerosis following bexarotene. Ann Clin Transl Neurol. Oct 2022;9(10):1626–1642. doi:10.1002/acn3.51662

76. De Stefano N, Matthews PM, Filippi M, et al. Evidence of early cortical atrophy in MS: relevance to white matter changes and disability. Neurology. Apr 8 2003;60(7):1157–62. doi:10.1212/01.wnl.0000055926.69643.03

77. Geurts JJ, Barkhof F. Grey matter pathology in multiple sclerosis. Lancet Neurol. Sep 2008;7(9):841–51. doi:10.1016/S1474-4422(08)70191-1

78. Lazzarotto A, Hamzaoui M, Tonietto M, et al. Time is myelin: early cortical myelin repair prevents atrophy and clinical progression in multiple sclerosis. Brain. Apr 4 2024;147(4):1331–1343. doi:10.1093/brain/awae024

79. Vercellino M, Masera S, Lorenzatti M, et al. Demyelination, inflammation, and neurodegeneration in multiple sclerosis deep gray matter. J Neuropathol Exp Neurol. May 2009;68(5):489–502. doi:10.1097/NEN.0b013e3181a19a5a

80. Morel A, Magnin M, Jeanmonod D. Multiarchitectonic and stereotactic atlas of the human thalamus. The Journal of Comparative Neurology. 1997;387(4):588–630. doi:10.1002/(sici)1096-9861(19971103)387:4<588::Aid-cne8>3.0.Co;2-z

81. Davies GR, Altmann DR, Rashid W, et al. Emergence of thalamic magnetization transfer ratio abnormality in early relapsing-remitting multiple sclerosis. Mult Scler. Jun 2005;11(3):276–81. doi:10.1191/1352458505ms1166oa

82. Al-Radaideh A, Athamneh I, Alabadi H, Hbahbih M. Deep gray matter changes in relapsing-remitting multiple sclerosis detected by multi-parametric, high-resolution magnetic resonance imaging (MRI). European radiology. Aug 26 2020;doi:10.1007/s00330-020-07199-5

83. Van Obberghen E, McHinda S, le Troter A, et al. Evaluation of the Sensitivity of Inhomogeneous Magnetization Transfer (ihMT) MRI for Multiple Sclerosis. AJNR Am J Neuroradiol. Apr 2018;39(4):634–641. doi:10.3174/ajnr.A5563

84. Yarnykh VL, Krutenkova EP, Aitmagambetova G, et al. Iron-Insensitive Quantitative Assessment of Subcortical Gray Matter Demyelination in Multiple Sclerosis Using the Macromolecular Proton Fraction. AJNR Am J Neuroradiol. Apr 2018;39(4):618–625. doi:10.3174/ajnr.A5542

85. Dutta R, Chang A, Doud MK, et al. Demyelination causes synaptic alterations in hippocampi from multiple sclerosis patients. Ann Neurol. Mar 2011;69(3):445–54. doi:10.1002/ana.22337

86. Xie Y, Zhang Y, Wu S, et al. Atrophy-Independent and Dependent Iron and Myelin Changes in Deep Gray Matter of Multiple Sclerosis: A Longitudinal Study Using chi- Separation Imaging. Acad Radiol. Jul 31 2024;doi:10.1016/j.acra.2024.07.031

87. Sicotte NL, Kern KC, Giesser BS, et al. Regional hippocampal atrophy in multiple sclerosis. Brain. Apr 2008;131(Pt 4):1134–41. doi:10.1093/brain/awn030

88. Planche V, Koubiyr I, Romero JE, et al. Regional hippocampal vulnerability in early multiple sclerosis: Dynamic pathological spreading from dentate gyrus to CA1. Hum Brain Mapp. Apr 2018;39(4):1814–1824. doi:10.1002/hbm.23970

89. Kohler W, Fischer M, Bublak P, et al. Information processing deficits as a driving force for memory impairment in MS: A cross--sectional study of memory functions and MRI in early and late stage MS. Mult Scler Relat Disord. Nov 2017;18:119–127. doi:10.1016/j.msard.2017.09.026

