## Supplement Material 1 for "Disability-relevant demyelination in multiple sclerosis: five-year evolution following diagnosis"

**Supplementary Material**

**Supplementary Table 1 Studies which have applied magnetization transfer saturation (MTsat) in people with multiple sclerosis (MS).**

| **Study** | **n** | **Disease duration, years**  **(MS only)** | **Age, years (MS only)** | **Sex (MS only)** | **Baseline EDSS** |
| --- | --- | --- | --- | --- | --- |
| Berg et al., (2022)^27^ | 11 RRMS; 1 SPMS; 1 CIS; 14 HC | 8 ± 5 (1 to 15) (range) | 34.3 ± 8.2 (24 to 46) (mean ± SD, range) | 8 F; 5 M | 1.1 ± 1.3 (0 to 4.5) (range) |
| Campbell et al., (2018)^31^ | 1 MS; 5 HC | - | - | - | - |
| Cagol et al., (2024)^42^ | 101 (active) RRMS MS; 82 (inactive) PMS; 73 HC | RRMS: 2.5 [0.6 to 9.3] (median [IQR])  PMS: 15.4 [8.0 to 25.0] | - | 110 F; 73 M | RRMS: 2.0 [1.5 to 2.5]  PMS: 5.0 [4.0 to 6.0] (median [IQR)] |
| Elkady et al., (2021)^28^ | 21 RRMS; 4 SPMS; 1 PPMS; 10 HC | 20.8 ± 9.6 (mean ± SD) | 50.2 ± 12.8 (mean ± SD) | 23 F; 3 M | 2.5 [0 to 6.5] (median, range) |
| Guillemin et al., (2024)^40^ | 19 RRMS/CIS; 19 HC | 1.79 ± 1.25 (mean ± SD) | 31.28 ± 5.21 (mean ± SD) | 13 F; 5 M | 1.75 [1.0 to 3.0] (median, range) |
| Lommers et al., (2019)^30^ | 15 RRMS; 7 SPMS; 14 PPMS; 36 HC | 13 [0.5 to 35] (median, range) | 45.69 ± 11.85 (mean ± SD) | 21 F; 15 M | 4.0 [1.0 to 6.5] (median, range) |
| Lommers et al., (2021)^29^ | 14 RRMS; 21 PMS; 36 HC | 13 [0.5 to 35] (median, range) | 46.2 ± 11.62 (mean ± SD) | 21 F; 14 M | 4.0 [1.0 to 6.0] (median, range) |
| Lema et al., (2017)^34^ | 128 RRMS; 6 SPMS | 3 [0 to 16] (median, range) | 39 [25 to 66] (median, range) | 107 F; 27 M | 3.5 [2.0 to 6.0] (median, range) |
| Longoni et al., (2023)^38^ | 11 ylPOMS; 18 HC | 2.5 [0.15 to 6.2] (mean, range) | 16.7 [14.2 to 18.2] (mean, range) | 7 F; 11 M | - |
| Mastantuono et al., (2026)^43^ | Cohort 1: 101 RRMS; 60 SPMS; 23 PPMS.  Cohort 2: 383 RRMS; 26 SPMS; 10 PPMS.  100 HC. | Cohort 1: active RRMS: 2.6 [0.7 to 9.4]; inactive PMS: 17.0 [8.0 to 26.0].  Cohort 2: active RRMS/SPMS: 2.8 [0.5 to 13.0]; inactive PMS: 12.2 [8.0 to 19.4] (median [IQR)]) | Cohort 1: active RRMS: 37.6 ± 10.9; inactive PMS: 58.3 ± 9.6  Cohort 2: active RRMS/SPMS: 40.3 ± 12.3; inactive PMS: 47.5 ± 12.0 (mean ± SD) | Cohort 1: active RRMS: 65 F; 36 M. Inactive PMS: 46 F; 37 M.  Cohort 2: active RRMS/SPMS: 43 F; 19 M. Inactive PMS: 235 F; 122 M. | Cohort 1: active RRMS: 2.0 [1.5 to 2.5]. Inactive PMS:.4.5 [4.0 to 6.0]  Cohort 2: active RRMS/SPMS:.2.5 [1.5 to 3.0] Inactive PMS: 2.5 [1.5 to 3.5]. (median [IQR)]. |
| Rahmanzadeh et al., (2022)^36^ | 92 (active) RRMS; 58 (inactive) PMS | - | 45 ± 15 (mean ± SD) | 92 F; 58 M | 3.65 [0 to 8.0] (median [range]) |
| Saccenti et al., (2020)^32^ | 21 RRMS | 8.7 ± 6.5 (mean ± SD) | 37.9 ± 9.9 (mean ± SD) | 19 F; 2 M | 1.0 (0 to 2.0) (range) |
| Sanabria-Diaz et al., (2024)^41^ | 75 RRMS; 53 PMS | 10.52 ± 10.77 (mean ± SD) | 46.80 ± 14.61 (mean ± SD) | 85 F; 43 M | 3.0 [1.5 to 4.5] (median [IQR]) |
| Schlaeger et al., (2025)^45^ | 11 RRMS; 1 SPMS; 1CIS | 8 ± 5 (1 to 15) (range) | 34.3 ± 8.2 (23 to 46) (mean ± SD, range) | 11 F; 3 M | 1.1 ± 1.3 (0 to 4.5) (range) |
| Tagge et al., (2022)^33^ | 18 RRMS; 6 SPMS; 1 PPMS | 16.3 ±8.5 (mean ± SD, RRMS); 22.5 ±7.8 (SPMS); 38 ±0 (PPMS) | 47.8 ±10.7 (mean ± SD, RRMS); 57.5 ±7.3 (SPMS); 66 ±0 (PPMS) | 19 F; 6 M | RRMS: 2.0; SPMS: 4.5; PMS: 6.5 (median) |
| Thevakumaran et al., (2026)^46^ | 19 RRMS; 19 PMS; 21 HC | RRMS: 18.0 [7.5]; PMS: 30.0 [15.5] (median, IQR) | RRMS: 48.9 [10.0]; PMS: 61.5 [8.6] (mean, SD) | 26 F; 12 M | RRMS: 2.5 [1.8]; PMS: 6.0 [2.5] (median [IQR]) |
| Trang et al., (2024)^47^ | 22 RRMS; 5 SPMS; 77 HC | 15.9 [13.2] (median, IQR) | 50 ± 9.9 (mean ± SD) | 18 F; 9 M | 3.0 [2] (median [IQR]) |
| Vandeleene et al., (2023)^39^ | 11 RRMS; 6 PMS | 3.4 [0.3 to 28] (at baseline, median, range) | 36 [25 to 65] (median, range) | 7 F; 10 M | 2.5 [1.0 to 6.5] (median [range]) |
| Wright et al., (2026)^44^ | 27 RRMS; 3 PPMS | 12.28 ± 7.06 (years ± SD) | 47.5 ± 11.5 (mean ± SD) | 20 F; 10 M | 3.0 [1.5 to 4.0] (median, [IQR]) |
| York et al., (2021)^35^ | 73 RRMS | ≤ 0.5 years since diagnosis | 33 [28 to 45] (median, IQR) | 57 F; 16 M | 2.0 [1.5 to 3.0] (median [IQR]) |
| York et al., (2022)^37^ | 62 RRMS | 3.6 [0.2 to 33.2] (since first symptom onset, 2 missing); 68 days [7 to 171] since diagnosis, mean, range); | 37.6 [21.7 to 67.3] (median, range) | 48 F; 14 M | 2.0 [0 to 6.0] (median [range]) |

Abbreviations: RRMS: relapsing-remitting MS; SPMS: secondary progressive MS; PPMS: primary progressive MS; PMS: progressive MS; CIS: clinically isolated syndrome; HC; healthy controls; EDSS: expanded disability status scale; IQR: interquartile range; SD: standard deviation; F/M: female/male; ylPOMS: youth living with pediatric-onset MS.

**Supplementary Table 2 MRI acquisition parameters.** Table adapted from Meijboom, Wiseman, York at al.^52^

| **Sequence** | **3D T1-w MPRAGE** | **2D FLAIR PROPELLER** | **2D T2-weighted DE FSE** | **3D FLASH spoiled GRE (MT_Off_/MT_On_/MT_T1w_)** |
| --- | --- | --- | --- | --- |
| **Orientation** | sagittal | axial | axial | sagittal |
| **FOV (mm)** | 256 | 250 | 250 | 224 (SI) x 241 (AP) |
| **Acq. Mat. (mm)** | 256 | 256 | 384 | 160 x 172 |
| **No. of slices recon.** | 176 | 60 | 60 | 128 |
| **Voxel size (mm)** | 1 x 1 x 1 | 1 x 1 x 3 | 0.7 x 0.7 x 3 | 1.4 x 1.4 x 1.4 |
| **TE (ms)** | 2.26 | 120 | 9.6/96 | 1.54/  4.55/  8.49 |
| **Acc. factor (in-plane x slice)** | 2 x 1 | 2 x 1 | 3 x 1 | 2 x 1 |
| **TR (ms)** | 2500 | 9500 | 3630 | 30 (MT_Off_ & MT_On_) / 15 (MT_T1w_) |
| **TI (ms)** | 1100 | 2400 | - | - |
| **FA (°)** | 7 | 150 | 150 | 5 (MT_Off_ & MT_On_) / 18 (MT_T1w_) |

Abbreviations: acq. mat.: acquisition matrix; DE: dual echo; FA: (excitation) flip angle; FLAIR: fluid attenuated inversion recovery; FLASH: fast low angle shot; FOV: field of view; GRE: gradient echo; MT: magnetization transfer; no. of slices recon.: number of slices reconstructed; TE: echo time; acc.:acceleration; TI: inversion time; TR: repetition time; T1-w: T1-weighted; T2-w: T2-weighted; Orient.: orientation; SE: spin echo.

**Supplementary Table 3: Demographics for people with relapsing-remitting multiple sclerosis (pwRRMS) who took part in the extended microstructural MRI FutureMS sub-study at baseline and 1-year follow-up, and completed EDSS assessment at 5-year follow-up.** Participants were dichotomised according to Expanded Disability Status Scale (EDSS) progression from baseline. DMT: disease-modifying treatment; EDSS: Expanded Disability Status Scale; F/M: female/male; ICV: intracranial volume; IQR: interquartile range; SD: standard deviation; WML: white matter lesion.

|  | Five-year follow-up | | |
| --- | --- | --- | --- |
|  | No progression | Progression | Lost to follow-up |
| n (F:M) | 37 (27:10) | 19 (15:4) | 6 (6:0) |
| Baseline age in years, mean (SD) | 35.3 (9.5) | 42.7 (12.0) | 37.3 (16.8) |
| EDSS, median [range] | 2.0 [0.0 to 6.5] | 4.5 [2.0 to 7.0] | - |
| Change in EDSS from previous time point, median [range] | 0.0 [-2.0 to 0.5] | 1.5 [0.5 to 3.0] | - |
| DMT between time points, n (%) | 28 (75.7%) | 13 (68.4%) | - |
| Time between first symptom onset and baseline visit, in years, median [IQR] | 2.33 [4.33] | 4.33 [6.88] | 3.13 [9.5] |
| Days between time points, median [IQR] | 1495 (106) | 1475 (83.5) | - |
| WML volume (as % of ICV), mean (SD) [n missing] | 0.522 (0.63) [1] | 0.669 (0.67) | - |
| Change in WML volume (as % of ICV) from previous time point, mean absolute difference (SD) | 0.025 (0.16) | 0.057 (0.15) | - |
| Progression at one-year follow-up, n [% of total] | 9 [34.6%] | 4 [28.6%] | 10 [45.5%] |

**Supplementary Table 4: Test-retest agreement from healthy control data (n=11).** 95% of median tissue measurements in healthy subjects would be expected to fall within the Bland-Altman limits of agreement.

|  |  | **MTsat (%)** | ***T*_1_ (s)** |
| --- | --- | --- | --- |
| White matter | Mean [SD] | 3.80 [0.12] | 1.09 [0.06] |
|  | Difference (mean [SD]) | 0.04 [0.10] | -0.03 [0.06] |
|  | Bland-Altman limits of agreement | ±0.20 | ±0.12 |
| Cortical grey matter | Mean [SD] | 1.86 (0.04) | 1.68 (0.07) |
|  | Difference (mean [SD]) | 0.01 [0.02] | -0.01 (0.06) |
|  | Bland-Altman limits of agreement | ±0.05 | ±0.11 |

Abbreviations: MTsat: magnetization transfer saturation; SD: standard deviation.

**Supplementary Table 5: Microstructural imaging summary.** n: number of participants; SD: standard deviation; WML: white matter lesion; YR0/1/5: baseline/1-year/5-year follow-up.

| **Metric** | **Brain Tissue** | **Baseline** | | | **One-year follow-up** | | **Annualised Change**  **(YR0 to YR1)** | |  | **Five-year follow-up** | | | **YR1 to YR5 change** | | **Annualised Change**  **(YR0 to YR1)** | | **Annualised Change**  **(YR1 to YR5)** | |
| --- | --- | --- | --- | --- | --- | --- | --- | --- | --- | --- | --- | --- | --- | --- | --- | --- | --- | --- |
|  |  | **n** | **Mean** | **SD** | **Mean** | **SD** | **Mean** | **SD** |  | **n** | **Mean** | **SD** | **Mean** | **SD** | **Mean** | **SD** | **Mean** | **SD** |
| MTsat | NAWM | 62 | 3.802 | 0.170 | 3.766 | 0.181 | -0.036 | 0.120 |  | 40 | 3.779 | 0.174 | 0.018 | 0.144 | -0.032 | 0.116 | 0.004 | 0.036 |
|  | GM |  | 1.891 | 0.044 | 1.882 | 0.048 | -0.01 | 0.04 |  |  | 1.864 | 0.051 | -0.011 | 0.041 | -0.009 | 0.036 | -0.003 | 0.01 |
|  | WML |  | 2.777 | 0.375 | 2.835 | 0.375 | 0.058 | 0.171 |  |  | 2.854 | 0.421 | 0.019 | 0.199 | 0.04 | 0.109 | 0.005 | 0.05 |
|  | DGM |  | 2.195 | 0.082 | 2.176 | 0.091 | -0.019 | 0.082 |  |  | 2.191 | 0.108 | 0.026 | 0.106 | -0.017 | 0.077 | 0.006 | 0.027 |
|  | Thalamus |  | 2.444 | 0.119 | 2.418 | 0.116 | -0.027 | 0.115 |  |  | 2.427 | 0.135 | 0.015 | 0.132 | -0.023 | 0.098 | 0.004 | 0.033 |
|  | HC |  | 1.6 | 0.067 | 1.6 | 0.074 | 0 | 0.069 |  |  | 1.608 | 0.102 | 0.015 | 0.113 | 0.003 | 0.067 | 0.004 | 0.028 |
|  | BG |  | 2.104 | 0.081 | 2.084 | 0.091 | -0.02 | 0.074 |  |  | 2.092 | 0.099 | 0.023 | 0.093 | -0.018 | 0.079 | 0.006 | 0.023 |
|  | Amygdala |  | 1.769 | 0.076 | 1.765 | 0.084 | -0.003 | 0.073 |  |  | 1.749 | 0.082 | -0.007 | 0.107 | 0.008 | 0.081 | -0.002 | 0.027 |
| T_1_ | NAWM | 62 | 1.08 | 0.066 | 1.09 | 0.063 | 0.01 | 0.057 |  | 40 | 1.072 | 0.07 | -0.014 | 0.077 | 0.007 | 0.06 | -0.003 | 0.019 |
|  | GM |  | 1.673 | 0.074 | 1.684 | 0.07 | 0.011 | 0.07 |  |  | 1.682 | 0.081 | -0.003 | 0.078 | 0.004 | 0.071 | -0.001 | 0.02 |
|  | WML |  | 1.34 | 0.161 | 1.338 | 0.157 | -0.002 | 0.078 |  |  | 1.332 | 0.165 | -0.002 | 0.096 | 0.005 | 0.07 | -0.001 | 0.024 |
|  | DGM |  | 1.742 | 0.091 | 1.748 | 0.087 | 0.006 | 0.093 |  |  | 1.728 | 0.098 | -0.015 | 0.116 | -0.001 | 0.099 | -0.004 | 0.029 |
|  | Thalamus |  | 1.798 | 0.092 | 1.811 | 0.088 | 0.013 | 0.1 |  |  | 1.792 | 0.097 | -0.012 | 0.113 | 0.005 | 0.104 | -0.003 | 0.028 |
|  | HC |  | 2.264 | 0.114 | 2.273 | 0.103 | 0.009 | 0.111 |  |  | 2.256 | 0.13 | -0.017 | 0.129 | -0.002 | 0.112 | -0.004 | 0.032 |
|  | BG |  | 1.682 | 0.096 | 1.684 | 0.093 | 0.002 | 0.089 |  |  | 1.663 | 0.101 | -0.019 | 0.115 | -0.004 | 0.093 | -0.005 | 0.029 |
|  | Amygdala |  | 2.2 | 0.117 | 2.197 | 0.11 | -0.003 | 0.117 |  |  | 2.19 | 0.129 | -0.018 | 0.134 | -0.008 | 0.117 | -0.005 | 0.034 |

**Supplementary Table 6: Relationship between 1-to-5-year change in MTsat and disability progression in early relapsing-remitting multiple sclerosis.** Models included the 1-year MTsat, age, 1-to-5-year change in lesion load, sex, and dichotomised disease-modifying therapies (DMTs) between 1-year and 5-years.

| **Metric** | **Outcome** | **n** | **Brain Tissue** | **β** | **z-value** | **Wald test** | | | **Adjusted odds ratio^a^ [95% CI]** | **Log-likelihood ratio (versus null model)** | | | **Log-likelihood ratio (versus reference^b^ model)** | | | **Nagelkerke’s (pseudo)-R^2^** |
| --- | --- | --- | --- | --- | --- | --- | --- | --- | --- | --- | --- | --- | --- | --- | --- | --- |
|  |  |  |  |  |  | **χ^2^** | **p-value** | **FDR-corr. p-value** |  | **diff.** | **χ^2^** | **p-value** | **diff.** | **χ^2^** | **p-value** |  |
| 1-to-5-year change in MTsat | 1-to-5-year EDSS progression | 40 | NAWM | 1.78 | 0.53 | 0.285 | 0.593 | 0.791 | 1.293 [0.503 to 3.47] | -4.13 | 8.25 | 0.22 | -0.235 | 0.47 | 0.791 | 0.26 |
|  |  |  | WML | -2.38 | -0.93 | 0.858 | 0.354 | 0.791 | 0.622 [0.195 to 1.58] | -4.62 | 9.24 | 0.161 | -0.726 | 1.452 | 0.484 | 0.28 |
|  |  |  | GM | 2.11 | 0.2 | 0.0406 | 0.84 | 0.84 | 1.09 [0.464 to 2.61] | -4.04 | 8.07 | 0.233 | -0.143 | 0.287 | 0.866 | 0.25 |
|  |  |  | DGM | -3.68 | -0.7 | 0.483 | 0.487 | 0.791 | 0.676 [0.189 to 1.72] | -4.17 | 8.35 | 0.214 | -0.282 | 0.564 | 0.754 | 0.26 |
|  |  |  | Thalamus | -4.48 | -0.91 | 0.829 | 0.363 | 0.84 | 0.553 [0.126 to 1.66] | -4.78 | 9.57 | 0.144 | -0.893 | 1.786 | 0.409 | 0.29 |
|  |  |  | HC | -3.61 | -0.71 | 0.5 | 0.479 | 0.84 | 0.666 [0.165 to 1.66] | -5.05 | 10.1 | 0.121 | -1.157 | 2.314 | 0.314 | 0.31 |
|  |  |  | BG | -1.99 | -0.39 | 0.153 | 0.696 | 0.84 | 0.831 [0.296 to 2.01] | -4.03 | 8.07 | 0.233 | -0.143 | 0.286 | 0.867 | 0.25 |
|  |  |  | Amygdala | -1.47 | -0.3 | 0.0884 | 0.766 | 0.84 | 0.854 [0.261 to 2.36] | -4.84 | 9.69 | 0.138 | -0.953 | 1.906 | 0.386 | 0.3 |

^a^For z-normalised predictors. ^b^Including baseline age, 1-to-5-year change in lesion load, sex, and DMTs between 1-year and 5-years. **P* < 0.05, ***P* < 0.01, ****P* < 0.001. Abbreviations: CI: confidence interval; BG: basal ganglia; DGM: deep grey matter; FDR: False Discovery Rate; GM: grey matter; HC: hippocampus; MTsat: magnetization transfer saturation; NAWM: ‘normal-appearing’ white matter; WML: white matter lesions.

**Supplementary Table 7: Influence of inflammatory disease activity post-hoc analyses.** Inflammatory disease activity was defined as the presence of new or enlarging white matter lesions (WML) at one-year follow-up. Results are shown for the interaction term for the given microstructural metric and inflammatory disease activity group. Covariates included the baseline value of the given microstructural metric, age, change in WML load over one year, sex, and dichotomised initiation of disease-modifying therapies (DMTs) over the first year.

| **Metric** | **EDSS**  **progression timescale** | **n** | **Brain Tissue** | **Interaction term** | | | | | |
| --- | --- | --- | --- | --- | --- | --- | --- | --- | --- |
|  |  |  |  | **β** | **z-value** | **Wald test** | | | **Adjusted odds ratio^a^ [95% CI]** |
|  |  |  |  |  |  | **χ^2^** | **p-value** | **FDR-corr. p-value** |  |
| Baseline-to-1-year MTsat | Baseline-to-1-year follow-up | 62 | NAWM | 5.64 | 0.736 | 0.542 | 0.461 | 0.826 | 1.97 [0.353 to 14.1] |
|  |  |  | GM | 28.7 | 1.2 | 1.45 | 0.229 | 0.826 | 3.19 [0.579 to 28.4] |
|  |  |  | WML | 2.59 | 0.411 | 0.169 | 0.681 | 0.826 | 1.56 [0.199 to 16.1] |
|  |  |  | DGM | -2.06 | -0.22 | 0.0485 | 0.826 | 0.826 | 0.845 [0.192 to 4.15] |
|  |  |  | Thalamus | -1.54 | -0.229 | 0.0526 | 0.819 | 0.826 | 0.838 [0.182 to 4.12] |
|  |  |  | HC | -6.3 | -0.594 | 0.353 | 0.552 | 0.826 | 0.649 [0.144 to 2.68] |
|  |  |  | BG | 4.28 | 0.444 | 0.197 | 0.657 | 0.826 | 1.38 [0.357 to 6.36] |

^a^For z-normalised predictors. Abbreviations: β: estimate for interaction term; BG: basal ganglia; CI: confidence interval; DGM: deep grey matter; GM: grey matter; HC: hippocampus; MTsat: magnetization transfer saturation; NAWM: ‘normal-appearing’ white matter; WML: white matter lesions present at baseline.

**Supplementary Table 8: Relationship between one-year change in T_1_ and disability progression in early relapsing-remitting multiple sclerosis.** Models included baseline T_1_, age, change in lesion load over one year, sex, and dichotomised initiation of disease-modifying therapies (DMTs) over the first year.

| **Metric** | **EDSS**  **progress.timescale** | **n** | **Brain Tissue** | **β** | **z-value** | **Wald test** | | | **Adjusted odds ratio^a^ [95% CI]** | **Log-likelihood ratio (versus null model)** | | | **Log-likelihood ratio (versus reference^b^ model)** | | | **Nagelkerke’s (pseudo)-R^2^** |
| --- | --- | --- | --- | --- | --- | --- | --- | --- | --- | --- | --- | --- | --- | --- | --- | --- |
|  |  |  |  |  |  | **χ^2^** | **p-value** | **FDR-corr. p-value** |  | **diff.** | **χ^2^** | **p-value** | **diff.** | **χ^2^** | **p-value** |  |
| Baseline-to-1-year change in T_1_ | Baseline-to-1-year follow-up | 62 | NAWM | 7.47 | 1.26 | 1.6 | 0.206 | 0.387 | 1.53 [0.807 to 3.14] | -3.11 | 6.22 | 0.399 | -1.349 | 2.698 | 0.26 | 0.13 |
|  |  |  | WML | 5.98 | 1.44 | 2.08 | 0.149 | 0.387 | 1.59 [0.864 to 3.14] | -2.97 | 5.94 | 0.429 | -1.211 | 2.422 | 0.298 | 0.12 |
|  |  |  | GM | 7.04 | 1.36 | 1.84 | 0.175 | 0.387 | 1.636 [0.821 to 3.46] | -2.8 | 5.6 | 0.47 | -1.038 | 2.075 | 0.354 | 0.12 |
|  |  |  | DGM | 3.94 | 1.06 | 1.12 | 0.29 | 0.387 | 1.441 [0.743 to 2.98] | -2.35 | 4.71 | 0.582 | -0.593 | 1.186 | 0.553 | 0.1 |
|  |  |  | Thalamus | 4.55 | 1.22 | 1.48 | 0.223 | 0.387 | 1.576 [0.773 to 3.46] | -2.54 | 5.08 | 0.534 | -0.779 | 1.557 | 0.459 | 0.11 |
|  |  |  | HC | 3.79 | 1.14 | 1.29 | 0.255 | 0.387 | 1.524 [0.758 to 3.29] | -2.52 | 5.05 | 0.538 | -0.763 | 1.526 | 0.466 | 0.11 |
|  |  |  | BG | 3.51 | 0.94 | 0.885 | 0.347 | 0.397 | 1.365 [0.721 to 2.73] | -2.23 | 4.47 | 0.614 | -0.471 | 0.943 | 0.624 | 0.09 |
|  |  |  | Amygdala | 0.9 | 0.31 | 0.096 | 0.756 | 0.756 | 1.111 [0.568 to 2.19] | -1.85 | 3.69 | 0.718 | -0.086 | 0.171 | 0.918 | 0.08 |
|  |  |  | Prolong. T_1_ WML voxels | 0.01 | 1.87 | 3.48 | 0.062 | 0.062 | 2.918 [1.18 to 12.7] | -5.16 | 10.31 | 0.112 | -3.396 | 6.792 | 0.034 | 0.21 |
|  | 1-year to 5-year follow-up | 56 | NAWM | -3.08 | -0.47 | 0.22 | 0.639 | 0.939 | 0.835 [0.377 to 1.73] | -3.71 | 7.43 | 0.283 | -0.113 | 0.226 | 0.893 | 0.17 |
|  |  |  | WML | 0.86 | 0.2 | 0.0398 | 0.842 | 0.939 | 1.07 [0.538 to 2.09] | -3.9 | 7.79 | 0.254 | -0.295 | 0.591 | 0.744 | 0.18 |
|  |  |  | GM | -1.83 | -0.35 | 0.121 | 0.728 | 0.939 | 0.875 [0.409 to 1.89] | -3.9 | 7.8 | 0.253 | -0.298 | 0.596 | 0.742 | 0.18 |
|  |  |  | DGM | -1.24 | -0.3 | 0.0916 | 0.762 | 0.939 | 0.889 [0.404 to 1.90] | -3.65 | 7.29 | 0.295 | -0.046 | 0.093 | 0.955 | 0.17 |
|  |  |  | Thalamus | -2.7 | -0.69 | 0.47 | 0.493 | 0.939 | 0.756 [0.33 to 1.67] | -3.89 | 7.77 | 0.255 | -0.285 | 0.57 | 0.752 | 0.18 |
|  |  |  | HC | -0.33 | -0.1 | 0.010 | 0.921 | 0.939 | 0.964 [0.461 to 2.07] | -4.04 | 8.08 | 0.233 | -0.437 | 0.875 | 0.646 | 0.19 |
|  |  |  | BG | -0.31 | -0.08 | 0.006 | 0.939 | 0.939 | 0.972 [0.458 to 2.02] | -3.6 | 7.21 | 0.302 | -0.004 | 0.008 | 0.996 | 0.17 |
|  |  |  | Amygdala | -1.05 | -0.34 | 0.116 | 0.734 | 0.939 | 0.884 [0.424 to 1.80] | -3.66 | 7.32 | 0.292 | -0.061 | 0.121 | 0.941 | 0.17 |
|  |  |  | Prolong. T_1_ WML voxels | 0 | -0.13 | 0.0164 | 0.898 | 0.898 | 0.943 [0.39 to 2.83] | -4.45 | 8.89 | 0.18 | -0.845 | 1.69 | 0.43 | 0.2 |

^a^For z-normalised predictors. ^b^Including age, change in lesion load over one year, sex, and initiation of DMTs. **P* < 0.05, ***P* < 0.01, ****P* < 0.001. Abbreviations: CI: confidence interval; BG: basal ganglia; DGM: deep grey matter; FDR: False Discovery Rate; GM: grey matter; HC: hippocampus; NAWM: ‘normal-appearing’ white matter; prolong. T_1_ WML voxels: number of WML voxels with T_1_ ≥2.00 seconds; WML: white matter lesions.

**Supplementary Table 9: Relationship between 1-to-5-year change in T_1_ and disability progression in early relapsing-remitting multiple sclerosis.** Models included the 1-year T_1_, age, 1-to-5-year change in lesion load, sex, and dichotomised disease-modifying therapies (DMTs) between 1-year and 5-years.

| **Metric** | **EDSS**  **progression timescale** | **n** | **Brain Tissue** | **β** | **z-value** | **Wald test** | | | **Adjusted odds ratio^a^ [95% CI]** | **Log-likelihood ratio (versus null model)** | | | **Log-likelihood ratio (versus reference^b^ model)** | | | **Nagelkerke’s (pseudo)-R^2^** |
| --- | --- | --- | --- | --- | --- | --- | --- | --- | --- | --- | --- | --- | --- | --- | --- | --- |
|  |  |  |  |  |  | **χ^2^** | **p-value** | **FDR-corr. p-value** |  | **diff.** | **χ^2^** | **p-value** | **diff.** | **χ^2^** | **p-value** |  |
| 1-to-5-year change in T_1_ | 1-to-5-year follow-up | 40 | NAWM | 3.03 | 0.49 | 0.238 | 0.626 | 0.629 | 1.262 [0.465 to 3.23] | -4.09 | 8.17 | 0.226 | -0.194 | 0.387 | 0.824 | 0.25 |
|  |  |  | WML | 2.86 | 0.66 | 0.435 | 0.509 | 0.629 | 1.315 [0.555 to 3.02] | -5.28 | 10.56 | 0.103 | -1.388 | 2.775 | 0.25 | 0.32 |
|  |  |  | GM | 2.82 | 0.48 | 0.234 | 0.629 | 0.629 | 1.247 [0.499 to 3.14] | -4.74 | 9.49 | 0.148 | -0.852 | 1.704 | 0.427 | 0.29 |
|  |  |  | DGM | 4.99 | 1.09 | 1.19 | 0.275 | 0.629 | 1.783 [0.615 to 5.31] | -5.39 | 10.78 | 0.095 | -1.499 | 2.999 | 0.223 | 0.33 |
|  |  |  | Thalamus | 3.22 | 0.69 | 0.481 | 0.488 | 0.629 | 1.438 [0.481 to 4.06] | -4.38 | 8.76 | 0.188 | -0.488 | 0.977 | 0.614 | 0.27 |
|  |  |  | HC | 6.29 | 1.64 | 2.7 | 0.101 | 0.629 | 2.246 [0.892 to 6.58] | -7.56 | 15.12 | 0.019 | -3.666 | 7.332 | 0.026 | 0.43 |
|  |  |  | BG | 5.31 | 1.15 | 1.32 | 0.25 | 0.629 | 1.842 [0.641 to 5.58] | -5.84 | 11.69 | 0.069 | -1.951 | 3.902 | 0.142 | 0.35 |
|  |  |  | Amygdala | 3.12 | 0.87 | 0.759 | 0.383 | 0.629 | 1.521 [0.555 to 4.1] | -5.37 | 10.75 | 0.097 | -1.481 | 2.962 | 0.227 | 0.32 |
|  |  |  | Prolong. T_1_ WML voxels | 0 | 0.74 | 0.547 | 0.46 | 0.46 | 1.424 [0.582 to 4.07] | -4.2 | 8.41 | 0.21 | -0.313 | 0.626 | 0.731 | 0.26 |

^a^For z-normalised predictors. ^b^Including baseline age, 1-to-5-year change in lesion load, sex, and DMTs between 1-year and 5-years. **P* < 0.05, ***P* < 0.01, ****P* < 0.001. Abbreviations: CI: confidence interval; BG: basal ganglia; DGM: deep grey matter; FDR: False Discovery Rate; GM: grey matter; HC: hippocampus; NAWM: ‘normal-appearing’ white matter; prolong. T_1_ WML voxels: number of WML voxels with T_1_ ≥2.00 seconds; WML: white matter lesions.

**Supplementary Table 10: Relationship between baseline T_1_ and 5-year disability progression in early relapsing-remitting multiple sclerosis.** Models included baseline T_1_, age, lesion load, and sex.

| **Metric** | **EDSS**  **progression timescale** | **n** | **Brain Tissue** | **β** | **z-value** | **Wald test** | | | **Adjusted odds ratio^a^ [95% CI]** | **Log-likelihood ratio (versus null model)** | | | **Log-likelihood ratio (versus reference^b^ model)** | | | **Nagelkerke’s (pseudo)-R^2^** |
| --- | --- | --- | --- | --- | --- | --- | --- | --- | --- | --- | --- | --- | --- | --- | --- | --- |
|  |  |  |  |  |  | **χ^2^** | **p-value** | **FDR-corr. p-value** |  | **diff.** | **χ^2^** | **p-value** | **diff.** | **χ^2^** | **p-value** |  |
| Baseline T_1_ | Baseline-to-5-year follow-up | 56 | NAWM | 1.63 | 0.35 | 0.121 | 0.728 | 0.885 | 1.111 [0.61 to 2.06] | -3.3 | 6.6 | 0.158 | -0.061 | 0.121 | 0.728 | 0.15 |
|  |  |  | WML | 0.86 | 0.49 | 0.235 | 0.628 | 0.885 | 1.15 [0.65 to 2.10] | -3.36 | 6.72 | 0.151 | -0.119 | 0.237 | 0.626 | 0.15 |
|  |  |  | GM | -0.94 | -0.23 | 0.0546 | 0.815 | 0.885 | 0.934 [0.519 to 1.66] | -3.27 | 6.54 | 0.162 | -0.027 | 0.055 | 0.815 | 0.15 |
|  |  |  | DGM | 0.46 | 0.14 | 0.021 | 0.885 | 0.885 | 1.042 [0.587 to 1.85] | -3.25 | 6.5 | 0.165 | -0.011 | 0.021 | 0.885 | 0.15 |
|  |  |  | Thalamus | -1.89 | -0.56 | 0.309 | 0.578 | 0.885 | 0.841 [0.442 to 1.53] | -3.4 | 6.8 | 0.147 | -0.157 | 0.315 | 0.575 | 0.15 |
|  |  |  | HC | 2.44 | 0.92 | 0.841 | 0.359 | 0.885 | 1.313 [0.752 to 2.52] | -3.69 | 7.38 | 0.117 | -0.449 | 0.897 | 0.344 | 0.16 |
|  |  |  | BG | 2.04 | 0.68 | 0.461 | 0.497 | 0.885 | 1.214 [0.696 to 2.19] | -3.48 | 6.95 | 0.138 | -0.234 | 0.468 | 0.494 | 0.16 |
|  |  |  | Amygdala | 0.53 | 0.22 | 0.0478 | 0.827 | 0.885 | 1.063 [0.606 to 1.89] | -3.27 | 6.53 | 0.163 | -0.024 | 0.048 | 0.827 | 0.15 |
|  |  |  | Prolong. T_1_ WML voxels | 0 | -1.26 | 1.58 | 0.209 | 0.209 | 0.567 [0.207 to 1.29] | -4.13 | 8.27 | 0.082 | -0.892 | 1.783 | 0.182 | 0.18 |

^a^For z-normalised predictors. ^b^Including baseline age, baseline lesion load and sex. **P* < 0.05, ***P* < 0.01, ****P* < 0.001. Abbreviations: CI: confidence interval; BG: basal ganglia; DGM: deep grey matter; FDR: False Discovery Rate; GM: grey matter; HC: hippocampus; NAWM: ‘normal-appearing’ white matter; prolong. T_1_ WML voxels: number of WML voxels with T_1_ ≥2.00 seconds; WML: white matter lesions.

**Supplementary Table 11: Relationship between baseline MTsat and 5-year disability progression in early relapsing-remitting multiple sclerosis.** Models included baseline MTsat, age, lesion load, and sex.

| **Metric** | **EDSS**  **progression timescale** | **n** | **Brain Tissue** | **β** | **z-value** | **Wald test** | | | **Adjusted odds ratio^a^ [95% CI]** | **Log-likelihood ratio (versus null model)** | | | **Log-likelihood ratio (versus reference^b^ model)** | | | **Nagelkerke’s (pseudo)-R^2^** |
| --- | --- | --- | --- | --- | --- | --- | --- | --- | --- | --- | --- | --- | --- | --- | --- | --- |
|  |  |  |  |  |  | **χ^2^** | **p-value** | **FDR-corr. p-value** |  | **diff.** | **χ^2^** | **p-value** | **diff.** | **χ^2^** | **p-value** |  |
| Baseline MTsat | Baseline-to-5-year follow-up | 56 | NAWM | -1.1 | -0.57 | 0.326 | 0.568 | 0.82 | 0.829 [0.428 to 1.57] | -3.41 | 6.81 | 0.146 | -0.164 | 0.329 | 0.566 | 0.15 |
|  |  |  | WML | -0.41 | -0.52 | 0.269 | 0.604 | 0.82 | 0.855 [0.464 to 1.55] | -3.38 | 6.75 | 0.149 | -0.135 | 0.271 | 0.603 | 0.15 |
|  |  |  | GM | 1.57 | 0.23 | 0.0519 | 0.82 | 0.82 | 1.068 [0.606 to 1.91] | -3.27 | 6.54 | 0.163 | -0.026 | 0.052 | 0.82 | 0.15 |
|  |  |  | DGM | 1.22 | 0.34 | 0.117 | 0.732 | 0.82 | 1.109 [0.618 to 2.06] | -3.3 | 6.6 | 0.158 | -0.059 | 0.119 | 0.731 | 0.15 |
|  |  |  | Thalamus | 4.7 | 1.54 | 2.37 | 0.124 | 0.824 | 1.768 [0.895 to 3.91] | -4.57 | 9.13 | 0.058 | -1.325 | 2.65 | 0.104 | 0.2 |
|  |  |  | HC | -3.48 | -0.8 | 0.633 | 0.426 | 0.824 | 0.786 [0.413 to 1.41] | -3.57 | 7.14 | 0.129 | -0.329 | 0.657 | 0.418 | 0.16 |
|  |  |  | BG | -0.8 | -0.22 | 0.0496 | 0.824 | 0.824 | 0.936 [0.517 to 1.69] | -3.27 | 6.53 | 0.163 | -0.025 | 0.05 | 0.824 | 0.15 |
|  |  |  | Amygdala | -1.79 | -0.47 | 0.218 | 0.641 | 0.824 | 0.872 [0.474 to 1.55] | -3.35 | 6.7 | 0.152 | -0.11 | 0.22 | 0.639 | 0.15 |

^a^For z-normalised predictors. ^b^Including baseline age, baseline lesion load and sex. **P* < 0.05, ***P* < 0.01, ****P* < 0.001. Abbreviations: CI: confidence interval; BG: basal ganglia; DGM: deep grey matter; FDR: False Discovery Rate; GM: grey matter; HC: hippocampus; MTsat: magnetization transfer saturation; NAWM: ‘normal-appearing’ white matter; WML: white matter lesions.

**Supplementary Table 12: Microstructural imaging summary by progression group.** Groups were defined according to Expanded Disability Status Scale (EDSS) progression between baseline and 1-year only (early progressors), between 1-year and 5-years only (late progressors), or both (progressors). BG: basal ganglia; DGM: deep grey matter; GM: grey matter; HC: hippocampus; n: number of participants; NAWM: ‘normal-appearing’ white matter; SD: standard deviation; WML: white matter lesion; YR0/1/5: baseline/1-year/5-year follow-up.

| **Metric** | **Group** | **n** | **Brain Tissue** | **Baseline** | | **One-year follow-up** | | **Five-year follow-up** | | **YR1 to YR5 change** | | **Annualised Change**  **(YR0 to YR1)** | | **Annualised Change**  **(YR1 to YR5)** | |
| --- | --- | --- | --- | --- | --- | --- | --- | --- | --- | --- | --- | --- | --- | --- | --- |
|  |  |  |  | **Mean** | **SD** | **Mean** | **SD** | **Mean** | **SD** | **Mean** | **SD** | **Mean** | **SD** | **Mean** | **SD** |
| MTsat | Non-progressors | 17 | NAWM | 3.85 | 0.146 | 3.832 | 0.195 | 3.833 | 0.158 | 0.001 | 0.15 | -0.019 | 0.112 | 0 | 0.037 |
|  |  |  | GM | 1.88 | 0.026 | 1.878 | 0.041 | 1.865 | 0.037 | -0.014 | 0.037 | -0.002 | 0.037 | -0.003 | 0.009 |
|  |  |  | WML | 2.855 | 0.403 | 2.94 | 0.404 | 2.986 | 0.383 | 0.046 | 0.131 | 0.086 | 0.088 | 0.012 | 0.033 |
|  |  |  | DGM | 2.198 | 0.08 | 2.191 | 0.1 | 2.208 | 0.08 | 0.017 | 0.082 | -0.007 | 0.066 | 0.004 | 0.021 |
|  |  |  | Thalamus | 2.454 | 0.116 | 2.442 | 0.123 | 2.451 | 0.112 | 0.01 | 0.098 | -0.012 | 0.091 | 0.002 | 0.024 |
|  |  |  | HC | 1.597 | 0.054 | 1.62 | 0.079 | 1.606 | 0.077 | -0.014 | 0.067 | 0.023 | 0.054 | -0.004 | 0.017 |
|  |  |  | BG | 2.11 | 0.068 | 2.096 | 0.096 | 2.112 | 0.07 | 0.015 | 0.086 | -0.014 | 0.064 | 0.004 | 0.022 |
|  |  |  | Amygdala | 1.749 | 0.053 | 1.776 | 0.079 | 1.735 | 0.068 | -0.041 | 0.073 | 0.027 | 0.062 | -0.01 | 0.018 |
|  | Early progressors | 9 | NAWM | 3.682 | 0.195 | 3.65 | 0.19 | 3.694 | 0.202 | 0.044 | 0.168 | -0.032 | 0.098 | 0.011 | 0.042 |
|  |  |  | GM | 1.891 | 0.046 | 1.866 | 0.048 | 1.868 | 0.072 | 0.002 | 0.048 | -0.024 | 0.04 | 0 | 0.012 |
|  |  |  | WML | 2.731 | 0.375 | 2.726 | 0.355 | 2.818 | 0.52 | 0.092 | 0.28 | -0.005 | 0.08 | 0.023 | 0.07 |
|  |  |  | DGM | 2.144 | 0.085 | 2.12 | 0.084 | 2.192 | 0.175 | 0.071 | 0.163 | -0.024 | 0.073 | 0.018 | 0.041 |
|  |  |  | Thalamus | 2.4 | 0.142 | 2.353 | 0.145 | 2.444 | 0.206 | 0.091 | 0.202 | -0.047 | 0.114 | 0.023 | 0.051 |
|  |  |  | HC | 1.584 | 0.041 | 1.56 | 0.07 | 1.649 | 0.175 | 0.089 | 0.196 | -0.024 | 0.076 | 0.022 | 0.049 |
|  |  |  | BG | 2.049 | 0.067 | 2.035 | 0.072 | 2.079 | 0.135 | 0.045 | 0.112 | -0.014 | 0.064 | 0.011 | 0.028 |
|  |  |  | Amygdala | 1.723 | 0.061 | 1.74 | 0.088 | 1.778 | 0.122 | 0.038 | 0.158 | 0.017 | 0.063 | 0.01 | 0.04 |
|  | Late progressors | 10 | NAWM | 3.776 | 0.165 | 3.762 | 0.146 | 3.757 | 0.179 | -0.005 | 0.085 | -0.014 | 0.084 | -0.001 | 0.021 |
|  |  |  | GM | 1.89 | 0.037 | 1.88 | 0.042 | 1.866 | 0.061 | -0.014 | 0.044 | -0.01 | 0.036 | -0.004 | 0.011 |
|  |  |  | WML | 2.766 | 0.356 | 2.792 | 0.373 | 2.75 | 0.339 | -0.041 | 0.121 | 0.025 | 0.135 | -0.01 | 0.03 |
|  |  |  | DGM | 2.171 | 0.085 | 2.171 | 0.115 | 2.151 | 0.085 | -0.021 | 0.073 | 0 | 0.089 | -0.005 | 0.018 |
|  |  |  | Thalamus | 2.41 | 0.102 | 2.417 | 0.139 | 2.37 | 0.109 | -0.048 | 0.097 | 0.008 | 0.102 | -0.012 | 0.024 |
|  |  |  | HC | 1.586 | 0.071 | 1.593 | 0.055 | 1.586 | 0.064 | -0.008 | 0.056 | 0.007 | 0.072 | -0.002 | 0.014 |
|  |  |  | BG | 2.065 | 0.1 | 2.067 | 0.119 | 2.052 | 0.102 | -0.015 | 0.066 | 0.001 | 0.087 | -0.004 | 0.016 |
|  |  |  | Amygdala | 1.757 | 0.056 | 1.764 | 0.065 | 1.736 | 0.054 | -0.027 | 0.071 | 0.006 | 0.08 | -0.007 | 0.018 |
|  | Progressors | 4 | NAWM | 3.84 | 0.169 | 3.705 | 0.204 | 3.792 | 0.12 | 0.087 | 0.204 | -0.135 | 0.211 | 0.022 | 0.051 |
|  |  |  | GM | 1.875 | 0.065 | 1.87 | 0.062 | 1.848 | 0.022 | -0.022 | 0.04 | -0.006 | 0.027 | -0.005 | 0.01 |
|  |  |  | WML | 2.755 | 0.308 | 2.743 | 0.197 | 2.633 | 0.498 | -0.109 | 0.343 | -0.012 | 0.143 | -0.027 | 0.086 |
|  |  |  | DGM | 2.224 | 0.067 | 2.142 | 0.103 | 2.214 | 0.077 | 0.072 | 0.083 | -0.082 | 0.091 | 0.018 | 0.021 |
|  |  |  | Thalamus | 2.5 | 0.101 | 2.409 | 0.113 | 2.43 | 0.079 | 0.02 | 0.099 | -0.09 | 0.067 | 0.005 | 0.025 |
|  |  |  | HC | 1.578 | 0.023 | 1.544 | 0.069 | 1.578 | 0.038 | 0.034 | 0.072 | -0.035 | 0.075 | 0.009 | 0.018 |
|  |  |  | BG | 2.136 | 0.073 | 2.042 | 0.134 | 2.141 | 0.109 | 0.099 | 0.112 | -0.094 | 0.13 | 0.025 | 0.028 |
|  |  |  | Amygdala | 1.766 | 0.085 | 1.681 | 0.14 | 1.768 | 0.091 | 0.088 | 0.111 | -0.085 | 0.144 | 0.022 | 0.028 |

**Supplementary Results**

**Primary Analyses**

**Associations with one-year disability progression**

**Normal-appearing white matter**

Data points for three participants (13.0%) with one-year progression and one (2.6%) without progression fell below the lower test-retest limit, and two participants (5.1%) without one-year progression exceeded the upper limit (Supplementary Table 4).

**White matter lesions**

Data points for all participants with one-year progression lay within white matter limits of agreement (Supplementary Table 4), with the exception of two participants (8.7%) who fell below the lower test-retest limit and one participant who exceed the upper limit (4.3%,). Six (15.4%) pwRRMS without one-year progression showed an increase in WML MTsat which exceeded the upper test-retest limit.

**Grey matter**

Of those participants with one-year progression, five (21.7%) data points fell below the lower test-retest limit for cortical GM MTsat and one (4.3%) exceeded the upper limit versus four (10.3%) below and three (7.7%) above for participants without progression (Supplementary Table 4).
