## Supplement Material 2 for "Disability-relevant demyelination in multiple sclerosis: five-year evolution following diagnosis"

**Diffusion tensor imaging (DTI) results**

| **Dependent variable: EDSS progression (YR0 to YR1)** | | | | | | | | | | |
| --- | --- | --- | --- | --- | --- | --- | --- | --- | --- | --- |
| **N** | **Metric** | **ROI** | **Beta** | ***z*-value** | **Wald χ^2^** | **Wald p-value** | **FDR-corrected p-value** | **Odds ratio (OR)** | **OR 2.5% confidence interval** | **OR 97.5% confidence interval** |
| 52 | YR0 DTI RD | WML | -230.64 | -0.04 | 0.00199 | 0.964 | 0.995 | 0.984 | 0.487 | 1.99 |
|  |  | NAWM | 1637.88 | 0.12 | 0.0142 | 0.905 | 0.98 | 1.04 | 0.537 | 2.02 |
|  |  | GM | -7962.8 | -0.28 | 0.081 | 0.776 | 0.98 | 0.916 | 0.495 | 1.69 |
|  |  | DGM | 3893.46 | 0.14 | 0.0193 | 0.89 | 0.98 | 1.049 | 0.542 | 2.14 |
|  |  | thalamus | -28823.2 | -1.14 | 1.3 | 0.254 | 0.98 | 0.691 | 0.355 | 1.29 |
|  |  | HC | 9817.11 | 0.59 | 0.348 | 0.555 | 0.98 | 1.208 | 0.642 | 2.3 |
|  |  | BG | 14812.56 | 0.59 | 0.354 | 0.552 | 0.98 | 1.263 | 0.603 | 2.85 |
|  |  | amygdala | 32787.45 | 1.61 | 2.58 | 0.108 | 0.98 | 1.742 | 0.904 | 3.58 |
|  | YR0 DTI AD | WML | 3685.47 | 1.08 | 1.17 | 0.279 | 0.98 | 1.415 | 0.763 | 2.74 |
|  |  | NAWM | 10274.84 | 0.78 | 0.613 | 0.434 | 0.98 | 1.31 | 0.666 | 2.63 |
|  |  | GM | 10198.97 | 0.31 | 0.0987 | 0.753 | 0.98 | 1.105 | 0.6 | 2.14 |
|  |  | DGM | 20803.52 | 0.93 | 0.861 | 0.353 | 0.98 | 1.431 | 0.698 | 3.23 |
|  |  | thalamus | 24245.83 | 1.12 | 1.26 | 0.261 | 0.98 | 1.525 | 0.755 | 3.39 |
|  |  | HC | 12164.13 | 0.79 | 0.619 | 0.432 | 0.98 | 1.283 | 0.692 | 2.48 |
|  |  | BG | 8291.01 | 0.39 | 0.15 | 0.698 | 0.98 | 1.147 | 0.582 | 2.4 |
|  |  | amygdala | 2885.25 | 0.25 | 0.0638 | 0.801 | 0.98 | 1.079 | 0.596 | 2.02 |
|  | YR0 DTI FA | WML | 8.79 | 1.32 | 1.74 | 0.187 | 0.98 | 1.575 | 0.832 | 3.34 |
|  |  | NAWM | 2.48 | 0.15 | 0.0214 | 0.884 | 0.98 | 1.049 | 0.554 | 2.02 |
|  |  | GM | 7.02 | 0.16 | 0.0249 | 0.875 | 0.98 | 1.051 | 0.569 | 1.98 |
|  |  | DGM | 4.22 | 0.21 | 0.044 | 0.834 | 0.98 | 1.073 | 0.558 | 2.1 |
|  |  | thalamus | 59.23 | 2.29 | 5.26 | 0.022 | 0.704 | 2.376 | 1.2 | 5.47 |
|  |  | HC | -13.11 | -0.4 | 0.163 | 0.686 | 0.98 | 0.876 | 0.455 | 1.69 |
|  |  | BG | -2.85 | -0.1 | 0.0104 | 0.919 | 0.98 | 0.956 | 0.404 | 2.35 |
|  |  | amygdala | -9.34 | -0.74 | 0.555 | 0.456 | 0.98 | 0.792 | 0.416 | 1.47 |
|  | YR0 DTI MD | WML | 2461.65 | 0.44 | 0.191 | 0.662 | 0.98 | 1.171 | 0.582 | 2.48 |
|  |  | NAWM | 6345.71 | 0.43 | 0.187 | 0.665 | 0.98 | 1.158 | 0.595 | 2.29 |
|  |  | GM | 155.92 | 0 | 2.43e-05 | 0.996 | 0.996 | 1.002 | 0.544 | 1.9 |
|  |  | DGM | 10890.39 | 0.35 | 0.125 | 0.724 | 0.98 | 1.136 | 0.591 | 2.49 |
|  |  | thalamus | -10120.9 | -0.38 | 0.143 | 0.705 | 0.98 | 0.885 | 0.47 | 1.7 |
|  |  | HC | 12913.81 | 0.75 | 0.569 | 0.45 | 0.98 | 1.27 | 0.682 | 2.43 |
|  |  | BG | 16191.03 | 0.62 | 0.386 | 0.535 | 0.98 | 1.257 | 0.632 | 2.73 |
|  |  | amygdala | 36182.34 | 1.47 | 2.17 | 0.141 | 0.98 | 1.622 | 0.869 | 3.22 |
| 56 | YR0 to YR1 change in DTI RD | WML | -2203.64 | -0.24 | 0.0583 | 0.809 | 0.946 | 0.925 | 0.487 | 1.8 |
|  |  | NAWM | 62142.41 | 0.95 | 0.906 | 0.341 | 0.881 | 1.351 | 0.74 | 2.62 |
|  |  | GM | -90685.7 | -1.74 | 3.04 | 0.081 | 0.648 | 0.568 | 0.288 | 1.05 |
|  |  | DGM | -12444.7 | -0.22 | 0.0473 | 0.828 | 0.946 | 0.935 | 0.505 | 1.73 |
|  |  | thalamus | 40639.4 | 0.75 | 0.562 | 0.454 | 0.881 | 1.283 | 0.67 | 2.53 |
|  |  | HC | 8355 | 0.26 | 0.07 | 0.791 | 0.946 | 1.092 | 0.563 | 2.14 |
|  |  | BG | -82810.9 | -1.56 | 2.43 | 0.119 | 0.667 | 0.622 | 0.332 | 1.11 |
|  |  | amygdala | 18187.01 | 0.61 | 0.366 | 0.545 | 0.881 | 1.238 | 0.625 | 2.55 |
|  | YR0 to YR1 change in DTI AD | WML | 8921.49 | 0.91 | 0.831 | 0.362 | 0.881 | 1.316 | 0.74 | 2.5 |
|  |  | NAWM | -23852 | -0.48 | 0.233 | 0.629 | 0.881 | 0.859 | 0.452 | 1.59 |
|  |  | GM | -129970 | -2.08 | 4.34 | 0.037 | 0.648 | 0.468 | 0.216 | 0.92 |
|  |  | DGM | -73214.8 | -1.45 | 2.12 | 0.146 | 0.667 | 0.593 | 0.28 | 1.17 |
|  |  | thalamus | 4439.22 | 0.11 | 0.0132 | 0.909 | 0.961 | 1.037 | 0.555 | 1.96 |
|  |  | HC | -1257.6 | -0.05 | 0.00256 | 0.96 | 0.961 | 0.984 | 0.517 | 1.85 |
|  |  | BG | -37201.8 | -0.83 | 0.688 | 0.407 | 0.881 | 0.766 | 0.396 | 1.42 |
|  |  | amygdala | -9662.23 | -0.49 | 0.236 | 0.627 | 0.881 | 0.837 | 0.378 | 1.68 |
|  | YR0 to YR1 change in DTI FA | WML | 5.11 | 0.5 | 0.253 | 0.615 | 0.881 | 1.178 | 0.607 | 2.37 |
|  |  | NAWM | -73.97 | -1.22 | 1.48 | 0.223 | 0.714 | 0.69 | 0.366 | 1.23 |
|  |  | GM | 63.17 | 0.72 | 0.52 | 0.471 | 0.881 | 1.242 | 0.692 | 2.28 |
|  |  | DGM | -93.08 | -1.5 | 2.25 | 0.134 | 0.667 | 0.602 | 0.295 | 1.13 |
|  |  | thalamus | -156.08 | -1.93 | 3.71 | 0.054 | 0.648 | 0.472 | 0.201 | 0.951 |
|  |  | HC | -3.1 | -0.05 | 0.00234 | 0.961 | 0.961 | 0.986 | 0.548 | 1.76 |
|  |  | BG | -34.49 | -0.48 | 0.227 | 0.633 | 0.881 | 0.866 | 0.469 | 1.57 |
|  |  | amygdala | -41.81 | -1.36 | 1.85 | 0.174 | 0.696 | 0.665 | 0.357 | 1.19 |
|  | YR0 to YR1 change in DTI MD | WML | 10594.88 | 0.87 | 0.757 | 0.384 | 0.881 | 1.367 | 0.722 | 3.11 |
|  |  | NAWM | 30632.03 | 0.41 | 0.17 | 0.68 | 0.907 | 1.139 | 0.614 | 2.18 |
|  |  | GM | -108013 | -1.86 | 3.45 | 0.063 | 0.648 | 0.534 | 0.263 | 1.01 |
|  |  | DGM | -34267.9 | -0.56 | 0.319 | 0.572 | 0.881 | 0.834 | 0.436 | 1.57 |
|  |  | thalamus | 29494.69 | 0.52 | 0.268 | 0.605 | 0.881 | 1.191 | 0.616 | 2.37 |
|  |  | HC | 2981.19 | 0.09 | 0.00833 | 0.927 | 0.961 | 1.031 | 0.535 | 2 |
|  |  | BG | -66728.7 | -1.22 | 1.5 | 0.221 | 0.714 | 0.68 | 0.356 | 1.24 |
|  |  | amygdala | 10836.98 | 0.32 | 0.104 | 0.747 | 0.946 | 1.126 | 0.544 | 2.36 |
| *DTI: diffusion tensor imaging; AD: axial diffusivity; MD: mean diffusivity; RD: radial diffusivity; FA: fractional anisotropy; WML: white matter lesions; NAWM: normal-appearing white matter; GM: grey matter; DGM: deep grey matter; HC: hippocampus; BG: basal ganglia; FDR: false discovery rate; ROI: region of interest; OR: odds ratio.* | | | | | | | | | | |

| **Dependent variable: EDSS progression (YR1 to YR5)** | | | | | | | | | | |
| --- | --- | --- | --- | --- | --- | --- | --- | --- | --- | --- |
| **N** | **Metric** | **ROI** | **Beta** | ***z*-value** | **Wald χ^2^** | **Wald p-value** | **FDR-corrected p-value** | **Odds ratio (OR)** | **OR 2.5% confidence interval** | **OR 97.5% confidence interval** |
| 52 | YR0 to YR1 change in DTI RD | WML | 3482.53 | 0.32 | 0.103 | 0.748 | 0.934 | 1.131 | 0.554 | 2.68 |
|  |  | NAWM | 76637.4 | 0.98 | 0.957 | 0.328 | 0.934 | 1.455 | 0.716 | 3.33 |
|  |  | GM | -8197.27 | -0.16 | 0.0249 | 0.875 | 0.934 | 0.949 | 0.486 | 1.83 |
|  |  | DGM | -54321.5 | -0.81 | 0.66 | 0.417 | 0.934 | 0.744 | 0.354 | 1.51 |
|  |  | thalamus | -52557.3 | -0.79 | 0.63 | 0.427 | 0.934 | 0.72 | 0.303 | 1.58 |
|  |  | HC | -4508.77 | -0.13 | 0.0169 | 0.897 | 0.934 | 0.955 | 0.468 | 1.91 |
|  |  | BG | -22601.2 | -0.4 | 0.159 | 0.69 | 0.934 | 0.877 | 0.453 | 1.69 |
|  |  | amygdala | -5908.99 | -0.18 | 0.0337 | 0.854 | 0.934 | 0.932 | 0.434 | 2.03 |
|  | YR0 to YR1 change in DTI AD | WML | -8497.52 | -0.8 | 0.637 | 0.425 | 0.934 | 0.765 | 0.381 | 1.47 |
|  |  | NAWM | 82479.75 | 1.4 | 1.96 | 0.162 | 0.934 | 1.691 | 0.835 | 3.75 |
|  |  | GM | -24699.4 | -0.41 | 0.171 | 0.679 | 0.934 | 0.863 | 0.421 | 1.76 |
|  |  | DGM | -44475.5 | -0.8 | 0.636 | 0.425 | 0.934 | 0.72 | 0.306 | 1.58 |
|  |  | thalamus | -59857.3 | -1.31 | 1.71 | 0.191 | 0.934 | 0.603 | 0.271 | 1.26 |
|  |  | HC | 15771.07 | 0.58 | 0.342 | 0.559 | 0.934 | 1.214 | 0.632 | 2.38 |
|  |  | BG | 34415.69 | 0.67 | 0.455 | 0.5 | 0.934 | 1.282 | 0.623 | 2.73 |
|  |  | amygdala | 37010.2 | 1.23 | 1.5 | 0.22 | 0.934 | 2.007 | 0.726 | 6.79 |
|  | YR0 to YR1 change in DTI FA | WML | -7.59 | -0.55 | 0.298 | 0.585 | 0.934 | 0.782 | 0.272 | 1.68 |
|  |  | NAWM | -22.29 | -0.34 | 0.114 | 0.736 | 0.934 | 0.894 | 0.452 | 1.73 |
|  |  | GM | 54.98 | 0.56 | 0.316 | 0.574 | 0.934 | 1.21 | 0.626 | 2.43 |
|  |  | DGM | 13.23 | 0.21 | 0.0431 | 0.836 | 0.934 | 1.075 | 0.53 | 2.14 |
|  |  | thalamus | -132.33 | -1.42 | 2.02 | 0.155 | 0.934 | 0.535 | 0.201 | 1.17 |
|  |  | HC | -11.12 | -0.15 | 0.0225 | 0.881 | 0.934 | 0.951 | 0.483 | 1.83 |
|  |  | BG | 103.42 | 1.27 | 1.61 | 0.204 | 0.934 | 1.558 | 0.808 | 3.27 |
|  |  | amygdala | 15.15 | 0.43 | 0.188 | 0.665 | 0.934 | 1.16 | 0.589 | 2.33 |
|  | YR0 to YR1 change in DTI MD | WML | 1433.06 | 0.12 | 0.0144 | 0.905 | 0.934 | 1.043 | 0.538 | 2.34 |
|  |  | NAWM | 123759.1 | 1.29 | 1.67 | 0.197 | 0.934 | 1.703 | 0.805 | 4.18 |
|  |  | GM | -21574 | -0.37 | 0.14 | 0.709 | 0.934 | 0.879 | 0.439 | 1.74 |
|  |  | DGM | -34520.1 | -0.49 | 0.242 | 0.623 | 0.934 | 0.83 | 0.387 | 1.75 |
|  |  | thalamus | -66634.9 | -0.94 | 0.89 | 0.346 | 0.934 | 0.669 | 0.278 | 1.52 |
|  |  | HC | 20914.66 | 0.58 | 0.331 | 0.565 | 0.934 | 1.22 | 0.618 | 2.49 |
|  |  | BG | -281.84 | 0 | 2.26e-05 | 0.996 | 0.996 | 0.998 | 0.495 | 2 |
|  |  | amygdala | 18525.53 | 0.49 | 0.245 | 0.621 | 0.934 | 1.23 | 0.555 | 2.93 |
| 37 | YR1 to YR5 change in DTI RD | WML | -4529.05 | -0.27 | 0.0734 | 0.786 | 0.929 | 0.862 | 0.264 | 2.47 |
|  |  | NAWM | -35212.1 | -0.78 | 0.607 | 0.436 | 0.872 | 0.71 | 0.25 | 1.61 |
|  |  | GM | -93168.8 | -1.11 | 1.24 | 0.265 | 0.872 | 0.439 | 0.0806 | 1.7 |
|  |  | DGM | 44095.25 | 0.89 | 0.784 | 0.376 | 0.872 | 1.591 | 0.579 | 4.89 |
|  |  | thalamus | -25003.7 | -0.43 | 0.183 | 0.668 | 0.902 | 0.803 | 0.264 | 2.13 |
|  |  | HC | 64445.19 | 1.38 | 1.89 | 0.169 | 0.872 | 2.154 | 0.764 | 7.36 |
|  |  | BG | 58441.37 | 1.45 | 2.09 | 0.148 | 0.872 | 2.453 | 0.807 | 10.8 |
|  |  | amygdala | 4921.52 | 0.13 | 0.0158 | 0.9 | 0.929 | 1.056 | 0.444 | 2.54 |
|  | YR1 to YR5 change in DTI AD | WML | 9207.3 | 0.79 | 0.627 | 0.428 | 0.872 | 1.712 | 0.548 | 8.11 |
|  |  | NAWM | 127013 | 1.89 | 3.57 | 0.059 | 0.872 | 4.043 | 1.2 | 21.8 |
|  |  | GM | 23675.94 | 0.38 | 0.143 | 0.705 | 0.902 | 1.248 | 0.398 | 4.18 |
|  |  | DGM | 13430.34 | 0.26 | 0.0695 | 0.792 | 0.929 | 1.178 | 0.34 | 4.26 |
|  |  | thalamus | 10290.26 | 0.18 | 0.0311 | 0.86 | 0.929 | 1.097 | 0.403 | 3.36 |
|  |  | HC | 37306.38 | 0.96 | 0.926 | 0.336 | 0.872 | 1.646 | 0.632 | 5.14 |
|  |  | BG | 43717.71 | 1.02 | 1.04 | 0.309 | 0.872 | 1.886 | 0.569 | 7.27 |
|  |  | amygdala | -33067.4 | -1.02 | 1.04 | 0.309 | 0.872 | 0.626 | 0.236 | 1.51 |
|  | YR1 to YR5 change in DTI FA | WML | 14.83 | 0.61 | 0.366 | 0.545 | 0.902 | 1.499 | 0.492 | 6.96 |
|  |  | NAWM | 36.37 | 0.96 | 0.928 | 0.335 | 0.872 | 1.601 | 0.726 | 6.67 |
|  |  | GM | 147.21 | 1.31 | 1.72 | 0.19 | 0.872 | 1.867 | 0.772 | 5.29 |
|  |  | DGM | 1.49 | 0.02 | 0.000477 | 0.983 | 0.983 | 1.011 | 0.381 | 2.7 |
|  |  | thalamus | 49.2 | 0.71 | 0.509 | 0.476 | 0.889 | 1.374 | 0.579 | 3.52 |
|  |  | HC | 30.86 | 0.39 | 0.154 | 0.695 | 0.902 | 1.201 | 0.491 | 3.27 |
|  |  | BG | -77.72 | -0.88 | 0.776 | 0.378 | 0.872 | 0.619 | 0.197 | 1.76 |
|  |  | amygdala | -22.23 | -0.54 | 0.287 | 0.592 | 0.902 | 0.78 | 0.289 | 1.9 |
|  | YR1 to YR5 change in DTI MD | WML | 2336.15 | 0.15 | 0.0222 | 0.882 | 0.929 | 1.087 | 0.365 | 3.54 |
|  |  | NAWM | 72176.14 | 0.86 | 0.746 | 0.388 | 0.872 | 1.589 | 0.564 | 4.95 |
|  |  | GM | -51841.3 | -0.67 | 0.454 | 0.5 | 0.889 | 0.64 | 0.156 | 2.31 |
|  |  | DGM | 11328.09 | 0.2 | 0.0388 | 0.844 | 0.929 | 1.111 | 0.371 | 3.31 |
|  |  | thalamus | -24454.3 | -0.38 | 0.147 | 0.702 | 0.902 | 0.816 | 0.265 | 2.31 |
|  |  | HC | 93212.81 | 1.74 | 3.03 | 0.082 | 0.872 | 2.771 | 0.991 | 10.6 |
|  |  | BG | 47097.84 | 1.13 | 1.27 | 0.26 | 0.872 | 2.038 | 0.623 | 8.28 |
|  |  | amygdala | -19728.5 | -0.41 | 0.168 | 0.682 | 0.902 | 0.832 | 0.328 | 2 |
| *DTI: diffusion tensor imaging; AD: axial diffusivity; MD: mean diffusivity; RD: radial diffusivity; FA: fractional anisotropy; WML: white matter lesions; NAWM: normal-appearing white matter; GM: grey matter; DGM: deep grey matter; HC: hippocampus; BG: basal ganglia; FDR: false discovery rate; ROI: region of interest; OR: odds ratio.* | | | | | | | | | | |

**Neurite Orientation Dispersion and Density Imaging (NODDI) results**

| **Dependent variable: EDSS progression (YR0 to YR1)** | | | | | | | | | | |
| --- | --- | --- | --- | --- | --- | --- | --- | --- | --- | --- |
| **N** | **Metric** | **ROI** | **Beta** | ***z*-value** | **Wald χ^2^** | **Wald p-value** | **FDR-corrected p-value** | **Odds ratio (OR)** | **OR 2.5% confidence interval** | **OR 97.5% confidence interval** |
| 56 | YR0 to YR1 change in NODDI ficvf | WML | -6.68 | -0.53 | 0.279 | 0.597 | 0.796 | 0.834 | 0.396 | 1.6 |
|  |  | NAWM | -26.46 | -0.56 | 0.315 | 0.575 | 0.796 | 0.837 | 0.437 | 1.54 |
|  |  | GM | 70.67 | 1.17 | 1.38 | 0.24 | 0.796 | 1.462 | 0.79 | 2.86 |
|  |  | DGM | 11.19 | 0.22 | 0.0501 | 0.823 | 0.962 | 1.073 | 0.572 | 2.02 |
|  |  | thalamus | -1.94 | -0.04 | 0.00169 | 0.967 | 0.967 | 0.987 | 0.52 | 1.87 |
|  |  | HC | -2.09 | -0.05 | 0.00293 | 0.957 | 0.967 | 0.983 | 0.525 | 1.84 |
|  |  | BG | 36.87 | 0.82 | 0.675 | 0.411 | 0.796 | 1.273 | 0.717 | 2.31 |
|  |  | amygdala | -32.06 | -0.89 | 0.797 | 0.372 | 0.796 | 0.707 | 0.318 | 1.49 |
|  | YR0 to YR1 change in NODDI odi | WML | -14.41 | -0.63 | 0.393 | 0.531 | 0.796 | 0.822 | 0.413 | 1.53 |
|  |  | NAWM | 146.01 | 0.9 | 0.806 | 0.369 | 0.796 | 1.308 | 0.731 | 2.4 |
|  |  | GM | 31.79 | 0.57 | 0.321 | 0.571 | 0.796 | 1.204 | 0.636 | 2.34 |
|  |  | DGM | 66.43 | 1.3 | 1.69 | 0.194 | 0.776 | 1.616 | 0.804 | 3.51 |
|  |  | thalamus | 121.22 | 1.82 | 3.3 | 0.069 | 0.414 | 1.83 | 0.982 | 3.71 |
|  |  | HC | -3.86 | -0.16 | 0.026 | 0.872 | 0.962 | 0.955 | 0.536 | 1.71 |
|  |  | BG | -11.25 | -0.29 | 0.086 | 0.769 | 0.962 | 0.91 | 0.466 | 1.7 |
|  |  | amygdala | 24.15 | 1.41 | 1.98 | 0.16 | 0.768 | 1.62 | 0.842 | 3.33 |
|  | YR0 to YR1 change in NODDI fiso | WML | -16.67 | -0.58 | 0.341 | 0.559 | 0.796 | 0.827 | 0.433 | 1.58 |
|  |  | NAWM | -38.56 | -0.77 | 0.587 | 0.443 | 0.796 | 0.781 | 0.4 | 1.45 |
|  |  | GM | -67.57 | -1.93 | 3.73 | 0.053 | 0.414 | 0.459 | 0.195 | 0.972 |
|  |  | DGM | 54.57 | 1.1 | 1.2 | 0.273 | 0.796 | 1.445 | 0.758 | 2.88 |
|  |  | thalamus | 24.47 | 0.62 | 0.38 | 0.538 | 0.796 | 1.215 | 0.661 | 2.36 |
|  |  | HC | 21.96 | 2.27 | 5.14 | 0.023 | 0.276 | 2.857 | 1.24 | 7.84 |
|  |  | BG | -4.14 | -0.15 | 0.022 | 0.882 | 0.962 | 0.951 | 0.476 | 1.86 |
|  |  | amygdala | -17.66 | -2.3 | 5.28 | 0.022 | 0.276 | 0.09 | 0.00656 | 0.44 |
| *DTI: diffusion tensor imaging; AD: axial diffusivity; MD: mean diffusivity; RD: radial diffusivity; FA: fractional anisotropy; WML: white matter lesions; NAWM: normal-appearing white matter; GM: grey matter; DGM: deep grey matter; HC: hippocampus; BG: basal ganglia; FDR: false discovery rate; ROI: region of interest; OR: odds ratio.* | | | | | | | | | | |

| **Dependent variable: EDSS progression (YR1 to YR5)** | | | | | | | | | | |
| --- | --- | --- | --- | --- | --- | --- | --- | --- | --- | --- |
| **N** | **Metric** | **ROI** | **Beta** | ***z*-value** | **Wald χ^2^** | **Wald p-value** | **FDR-corrected p-value** | **Odds ratio (OR)** | **OR 2.5% confidence interval** | **OR 97.5% confidence interval** |
| 52 |  | WML | -2.67 | -0.42 | 0.178 | 0.673 | 0.85 | 0.865 | 0.428 | 1.7 |
|  |  | NAWM | -6.31 | -0.58 | 0.337 | 0.561 | 0.799 | 0.82 | 0.41 | 1.6 |
|  |  | GM | -5 | -0.14 | 0.0207 | 0.886 | 0.886 | 0.956 | 0.508 | 1.78 |
|  |  | DGM | -5.09 | -0.17 | 0.0302 | 0.862 | 0.886 | 0.943 | 0.464 | 1.78 |
|  |  | thalamus | 16.77 | 0.65 | 0.418 | 0.518 | 0.799 | 1.226 | 0.655 | 2.31 |
|  |  | HC | -23.18 | -1.02 | 1.05 | 0.306 | 0.67 | 0.718 | 0.369 | 1.35 |
|  |  | BG | -16.02 | -0.66 | 0.435 | 0.509 | 0.799 | 0.791 | 0.381 | 1.58 |
|  |  | amygdala | -49.78 | -1.86 | 3.45 | 0.063 | 0.45 | 0.526 | 0.253 | 1 |
|  |  | WML | -10.09 | -1.13 | 1.27 | 0.26 | 0.67 | 0.706 | 0.37 | 1.27 |
|  |  | NAWM | -62.96 | -1.08 | 1.18 | 0.278 | 0.67 | 0.675 | 0.32 | 1.35 |
|  |  | GM | -6.35 | -0.22 | 0.0481 | 0.826 | 0.886 | 0.934 | 0.497 | 1.72 |
|  |  | DGM | -7.01 | -0.47 | 0.219 | 0.64 | 0.85 | 0.856 | 0.437 | 1.63 |
|  |  | thalamus | -50.69 | -1.85 | 3.41 | 0.065 | 0.45 | 0.502 | 0.227 | 1 |
|  |  | HC | -3.16 | -0.25 | 0.0601 | 0.806 | 0.886 | 0.922 | 0.466 | 1.77 |
|  |  | BG | -8.9 | -0.57 | 0.33 | 0.566 | 0.799 | 0.78 | 0.32 | 1.79 |
|  |  | amygdala | 4.43 | 0.63 | 0.399 | 0.527 | 0.799 | 1.219 | 0.656 | 2.31 |
|  |  | WML | -11.23 | -0.68 | 0.458 | 0.498 | 0.799 | 0.774 | 0.347 | 1.59 |
|  |  | NAWM | -40.03 | -1.02 | 1.04 | 0.307 | 0.67 | 0.708 | 0.35 | 1.35 |
|  |  | GM | -25.99 | -1.21 | 1.46 | 0.227 | 0.67 | 0.619 | 0.269 | 1.32 |
|  |  | DGM | 74.29 | 1.56 | 2.45 | 0.118 | 0.472 | 2.332 | 0.871 | 7.31 |
|  |  | thalamus | 94.77 | 1.78 | 3.16 | 0.075 | 0.45 | 2.303 | 0.964 | 6.28 |
|  |  | HC | 2.2 | 0.31 | 0.093 | 0.76 | 0.886 | 1.112 | 0.56 | 2.26 |
|  |  | BG | 79.06 | 2.39 | 5.7 | 0.017 | 0.408 | 3.973 | 1.41 | 14.1 |
|  |  | amygdala | 4.32 | 1.64 | 2.69 | 0.101 | 0.472 | 1.85 | 0.952 | 4.28 |
| 52 | YR0 to YR1 change in NODDI ficvf | WML | -4.62 | -0.33 | 0.107 | 0.744 | 0.957 | 0.882 | 0.372 | 1.81 |
|  |  | NAWM | -27.76 | -0.52 | 0.271 | 0.603 | 0.957 | 0.83 | 0.397 | 1.65 |
|  |  | GM | 16.63 | 0.27 | 0.072 | 0.788 | 0.957 | 1.096 | 0.559 | 2.19 |
|  |  | DGM | 61.94 | 1.08 | 1.16 | 0.281 | 0.957 | 1.494 | 0.725 | 3.21 |
|  |  | thalamus | 52.7 | 0.94 | 0.878 | 0.349 | 0.957 | 1.443 | 0.684 | 3.27 |
|  |  | HC | 1.29 | 0.03 | 0.00091 | 0.976 | 0.976 | 1.01 | 0.522 | 1.99 |
|  |  | BG | 5.21 | 0.1 | 0.0109 | 0.917 | 0.957 | 1.036 | 0.527 | 2.01 |
|  |  | amygdala | -5.1 | -0.14 | 0.0187 | 0.891 | 0.957 | 0.945 | 0.41 | 2.13 |
|  | YR0 to YR1 change in NODDI odi | WML | 20.75 | 0.65 | 0.426 | 0.514 | 0.957 | 1.337 | 0.619 | 3.51 |
|  |  | NAWM | -49.43 | -0.28 | 0.0797 | 0.778 | 0.957 | 0.911 | 0.463 | 1.73 |
|  |  | GM | -27.32 | -0.45 | 0.2 | 0.655 | 0.957 | 0.851 | 0.414 | 1.74 |
|  |  | DGM | 25.84 | 0.49 | 0.241 | 0.624 | 0.957 | 1.21 | 0.571 | 2.7 |
|  |  | thalamus | 141.3 | 1.79 | 3.22 | 0.073 | 0.957 | 2.029 | 0.987 | 4.81 |
|  |  | HC | -13.87 | -0.54 | 0.295 | 0.587 | 0.957 | 0.843 | 0.442 | 1.57 |
|  |  | BG | -35.11 | -0.81 | 0.648 | 0.421 | 0.957 | 0.742 | 0.338 | 1.49 |
|  |  | amygdala | -16.05 | -0.8 | 0.644 | 0.422 | 0.957 | 0.723 | 0.306 | 1.54 |
|  | YR0 to YR1 change in NODDI fiso | WML | 24.85 | 0.77 | 0.59 | 0.442 | 0.957 | 1.332 | 0.656 | 2.89 |
|  |  | NAWM | 10.36 | 0.18 | 0.0332 | 0.856 | 0.957 | 1.067 | 0.514 | 2.14 |
|  |  | GM | -12.79 | -0.4 | 0.162 | 0.687 | 0.957 | 0.86 | 0.401 | 1.79 |
|  |  | DGM | 39.13 | 0.67 | 0.445 | 0.505 | 0.957 | 1.299 | 0.615 | 2.95 |
|  |  | thalamus | 46.5 | 0.82 | 0.672 | 0.412 | 0.957 | 1.401 | 0.659 | 3.42 |
|  |  | HC | -9.86 | -1.14 | 1.3 | 0.255 | 0.957 | 0.627 | 0.268 | 1.39 |
|  |  | BG | -4.84 | -0.15 | 0.0227 | 0.88 | 0.957 | 0.942 | 0.426 | 2.1 |
|  |  | amygdala | -11.38 | -1.32 | 1.74 | 0.187 | 0.957 | 0.218 | 0.0106 | 1.14 |
| 37 | YR1 to YR5 change in NODDI ficvf | WML | -20.16 | -0.99 | 0.985 | 0.321 | 0.583 | 0.542 | 0.144 | 1.72 |
|  |  | NAWM | -5.85 | -0.17 | 0.0273 | 0.869 | 0.907 | 0.93 | 0.376 | 2.25 |
|  |  | GM | -44.11 | -0.85 | 0.724 | 0.395 | 0.583 | 0.521 | 0.0546 | 1.41 |
|  |  | DGM | -115.01 | -1.42 | 2.02 | 0.155 | 0.583 | 0.34 | 0.0624 | 1.36 |
|  |  | thalamus | -46.36 | -0.72 | 0.523 | 0.469 | 0.592 | 0.663 | 0.201 | 2.01 |
|  |  | HC | -63.29 | -1.15 | 1.32 | 0.251 | 0.583 | 0.394 | 0.0531 | 1.17 |
|  |  | BG | -85.39 | -1.36 | 1.85 | 0.174 | 0.583 | 0.34 | 0.048 | 1.36 |
|  |  | amygdala | 58.56 | 1.09 | 1.19 | 0.276 | 0.583 | 1.732 | 0.692 | 5.35 |
|  | YR1 to YR5 change in NODDI odi | WML | -67.27 | -1.57 | 2.45 | 0.117 | 0.583 | 0.277 | 0.0434 | 1.09 |
|  |  | NAWM | -272.87 | -1.37 | 1.88 | 0.17 | 0.583 | 0.285 | 0.0352 | 1 |
|  |  | GM | -126.53 | -1.89 | 3.56 | 0.059 | 0.583 | 0.408 | 0.14 | 0.957 |
|  |  | DGM | 22.95 | 0.36 | 0.131 | 0.717 | 0.782 | 1.208 | 0.441 | 3.57 |
|  |  | thalamus | -96 | -1.14 | 1.31 | 0.253 | 0.583 | 0.577 | 0.203 | 1.43 |
|  |  | HC | -30.8 | -1 | 0.997 | 0.318 | 0.583 | 0.62 | 0.219 | 1.52 |
|  |  | BG | 19.82 | 0.41 | 0.17 | 0.68 | 0.777 | 1.23 | 0.465 | 3.59 |
|  |  | amygdala | 17.77 | 0.82 | 0.67 | 0.413 | 0.583 | 1.459 | 0.589 | 3.85 |
|  | YR1 to YR5 change in NODDI fiso | WML | -19.17 | -1.03 | 1.07 | 0.301 | 0.583 | 0.622 | 0.202 | 1.39 |
|  |  | NAWM | -32.26 | -0.68 | 0.463 | 0.496 | 0.595 | 0.737 | 0.279 | 1.75 |
|  |  | GM | -30.78 | -0.87 | 0.751 | 0.386 | 0.583 | 0.342 | 0.0181 | 1.18 |
|  |  | DGM | -48.41 | -0.91 | 0.821 | 0.365 | 0.583 | 0.555 | 0.0869 | 1.41 |
|  |  | thalamus | -32.46 | -0.75 | 0.564 | 0.453 | 0.592 | 0.696 | 0.166 | 1.57 |
|  |  | HC | 0.3 | 0.02 | 0.000463 | 0.983 | 0.983 | 1.013 | 0.289 | 3.28 |
|  |  | BG | -45.72 | -1.28 | 1.63 | 0.202 | 0.583 | 0.426 | 0.081 | 1.2 |
|  |  | amygdala | -14.94 | -1.08 | 1.18 | 0.278 | 0.583 | 0.186 | 0.00427 | 2.29 |
| *DTI: diffusion tensor imaging; AD: axial diffusivity; MD: mean diffusivity; RD: radial diffusivity; FA: fractional anisotropy; WML: white matter lesions; NAWM: normal-appearing white matter; GM: grey matter; DGM: deep grey matter; HC: hippocampus; BG: basal ganglia; FDR: false discovery rate; ROI: region of interest; OR: odds ratio.* | | | | | | | | | | |
