## Supplement Material 3 for "Disability-relevant demyelination in multiple sclerosis: five-year evolution following diagnosis"

**Results for blood biomarkers**

| **Dependent variable: YR0 to YR1 EDSS progression** | | | | | | | | | | |
| --- | --- | --- | --- | --- | --- | --- | --- | --- | --- | --- |
| **n** | **Blood**  **metric** | **MRI metric** | **ROI** | **Results for blood metric in multivariable model** | | | | | | **Likelihood Ratio Test (with vs without blood metric) p-value** |
|  |  |  |  | **Beta** | **z-value** | **p-value (uncorrected)** | **Odds ratio (OR)** | **OR 2.5% confidence interval** | **OR 97.5% confidence interval** |  |
| 59 | YR0 GFAP z-score | YR0 to YR1 change in MTsat | Amygdala | -0.08 | -0.3 | 0.796 | 0.92 | 0.491 | 1.72 | 0.794 |
|  |  |  | BG | -0.04 | -0.1 | 0.916 | 0.96 | 0.49 | 1.9 | 0.916 |
|  |  |  | DGM | -0.05 | -0.1 | 0.881 | 0.95 | 0.473 | 1.9 | 0.881 |
|  |  |  | GM | -0.04 | -0.1 | 0.902 | 0.96 | 0.477 | 1.92 | 0.902 |
|  |  |  | HC | -0.25 | -0.7 | 0.506 | 0.78 | 0.366 | 1.64 | 0.494 |
|  |  |  | Thalamus | -0.09 | -0.3 | 0.793 | 0.91 | 0.455 | 1.82 | 0.791 |
|  |  |  | NAWM | 0.02 | 0 | 0.963 | 1.02 | 0.49 | 2.11 | 0.963 |
|  |  |  | WML | -0.13 | -0.4 | 0.696 | 0.88 | 0.455 | 1.69 | 0.692 |
|  |  | YR0 to YR1 change in T1 | Amygdala | -0.15 | -0.5 | 0.634 | 0.86 | 0.461 | 1.6 | 0.63 |
|  |  |  | BG | -0.1 | -0.3 | 0.749 | 0.9 | 0.476 | 1.71 | 0.747 |
|  |  |  | DGM | -0.09 | -0.3 | 0.766 | 0.91 | 0.48 | 1.72 | 0.764 |
|  |  |  | GM | -0.11 | -0.3 | 0.73 | 0.89 | 0.472 | 1.69 | 0.727 |
|  |  |  | HC | -0.13 | -0.4 | 0.667 | 0.87 | 0.468 | 1.63 | 0.663 |
|  |  |  | Thalamus | -0.09 | -0.3 | 0.774 | 0.91 | 0.483 | 1.72 | 0.772 |
|  |  |  | NAWM | -0.03 | -0.1 | 0.926 | 0.97 | 0.51 | 1.84 | 0.926 |
|  |  |  | WML | -0.11 | -0.4 | 0.709 | 0.89 | 0.477 | 1.65 | 0.706 |
|  | YR0 EBNA1 antibodies (log) | YR0 to YR1 change in MTsat | Amygdala | -0.11 | -0.8 | 0.451 | 0.8 | 0.454 | 1.42 | 0.455 |
|  |  |  | BG | -0.14 | -0.9 | 0.352 | 0.76 | 0.425 | 1.36 | 0.351 |
|  |  |  | DGM | -0.13 | -0.9 | 0.383 | 0.77 | 0.428 | 1.38 | 0.381 |
|  |  |  | GM | -0.13 | -0.9 | 0.381 | 0.77 | 0.421 | 1.39 | 0.38 |
|  |  |  | HC | -0.13 | -0.9 | 0.388 | 0.77 | 0.418 | 1.4 | 0.395 |
|  |  |  | Thalamus | -0.14 | -1 | 0.341 | 0.75 | 0.42 | 1.35 | 0.34 |
|  |  |  | NAWM | -0.18 | -1.1 | 0.256 | 0.7 | 0.373 | 1.3 | 0.265 |
|  |  |  | WML | -0.19 | -1.2 | 0.221 | 0.69 | 0.381 | 1.25 | 0.215 |
|  |  | YR0 to YR1 change in T1 | Amygdala | -0.12 | -0.8 | 0.432 | 0.79 | 0.447 | 1.41 | 0.434 |
|  |  |  | BG | -0.1 | -0.7 | 0.476 | 0.82 | 0.465 | 1.43 | 0.478 |
|  |  |  | DGM | -0.1 | -0.7 | 0.486 | 0.82 | 0.467 | 1.44 | 0.488 |
|  |  |  | GM | -0.11 | -0.8 | 0.442 | 0.8 | 0.458 | 1.41 | 0.446 |
|  |  |  | HC | -0.09 | -0.6 | 0.544 | 0.84 | 0.476 | 1.48 | 0.547 |
|  |  |  | Thalamus | -0.1 | -0.7 | 0.498 | 0.82 | 0.468 | 1.45 | 0.5 |
|  |  |  | NAWM | -0.1 | -0.7 | 0.479 | 0.82 | 0.463 | 1.44 | 0.484 |
|  |  |  | WML | -0.14 | -1 | 0.32 | 0.75 | 0.424 | 1.32 | 0.32 |
|  | YR0 NfL z-score | YR0 to YR1 change in MTsat | Amygdala | 0.19 | 0.8 | 0.429 | 1.33 | 0.655 | 2.71 | 0.421 |
|  |  |  | BG | 0.2 | 0.8 | 0.427 | 1.34 | 0.653 | 2.73 | 0.423 |
|  |  |  | DGM | 0.18 | 0.7 | 0.466 | 1.31 | 0.635 | 2.7 | 0.46 |
|  |  |  | GM | 0.33 | 1.2 | 0.222 | 1.62 | 0.746 | 3.51 | 0.203 |
|  |  |  | HC | 0.15 | 0.6 | 0.556 | 1.25 | 0.593 | 2.64 | 0.552 |
|  |  |  | Thalamus | 0.2 | 0.8 | 0.425 | 1.34 | 0.653 | 2.75 | 0.418 |
|  |  |  | NAWM | 0.21 | 0.7 | 0.454 | 1.37 | 0.602 | 3.11 | 0.448 |
|  |  |  | WML | 0.3 | 1.1 | 0.261 | 1.56 | 0.717 | 3.4 | 0.249 |
|  |  | YR0 to YR1 change in T1 | Amygdala | 0.22 | 0.9 | 0.376 | 1.38 | 0.674 | 2.84 | 0.369 |
|  |  |  | BG | 0.26 | 1 | 0.307 | 1.46 | 0.705 | 3.04 | 0.297 |
|  |  |  | DGM | 0.27 | 1 | 0.295 | 1.48 | 0.709 | 3.11 | 0.282 |
|  |  |  | GM | 0.27 | 1 | 0.295 | 1.48 | 0.71 | 3.1 | 0.282 |
|  |  |  | HC | 0.24 | 1 | 0.328 | 1.43 | 0.696 | 2.95 | 0.317 |
|  |  |  | Thalamus | 0.28 | 1.1 | 0.283 | 1.51 | 0.713 | 3.19 | 0.267 |
|  |  |  | NAWM | 0.25 | 1 | 0.336 | 1.44 | 0.684 | 3.05 | 0.323 |
|  |  |  | WML | 0.37 | 1.3 | 0.19 | 1.72 | 0.763 | 3.9 | 0.167 |
| *WML: white matter lesions; NAWM: normal-appearing white matter; GM: grey matter; DGM: deep grey matter; HC: hippocampus; BG: basal ganglia; FDR: false discovery rate; ROI: region of interest; OR: odds ratio; YR0: Baseline (year 0); YR1: 1-year follow-up; MTsat: magnetisation transfer saturation; EDSS: Expanded Disability Status Scale.* | | | | | | | | | | |

| **Dependent variable: YR1 to YR5 EDSS progression** | | | | | | | | | | |
| --- | --- | --- | --- | --- | --- | --- | --- | --- | --- | --- |
| **n** | **Blood**  **metric** | **MRI metric** | **ROI** | **Results for blood metric in multivariable model** | | | | | | **Likelihood Ratio Test (with vs without blood metric) p-value** |
|  |  |  |  | **Beta** | **z-value** | **p-value (uncorrected)** | **Odds ratio (OR)** | **OR 2.5% confidence interval** | **OR 97.5% confidence interval** |  |
| 38 | YR0 GFAP z-score | YR1 to YR5 change in MTsat | Amygdala | 0.36 | 0.7 | 0.463 | 1.42 | 0.558 | 3.6 | 0.455 |
|  |  |  | BG | 0.36 | 0.7 | 0.485 | 1.42 | 0.529 | 3.82 | 0.479 |
|  |  |  | DGM | 0.3 | 0.6 | 0.559 | 1.34 | 0.504 | 3.55 | 0.555 |
|  |  |  | GM | 0.19 | 0.4 | 0.7 | 1.21 | 0.462 | 3.16 | 0.699 |
|  |  |  | HC | 0.15 | 0.3 | 0.77 | 1.15 | 0.443 | 3 | 0.768 |
|  |  |  | Thalamus | 0.19 | 0.3 | 0.727 | 1.2 | 0.435 | 3.3 | 0.726 |
|  |  |  | NAWM | 0.2 | 0.4 | 0.682 | 1.22 | 0.473 | 3.14 | 0.68 |
|  |  |  | WML | 0.16 | 0.3 | 0.749 | 1.17 | 0.446 | 3.07 | 0.747 |
|  |  | YR1 to YR5 change in T1 | Amygdala | 0.33 | 0.7 | 0.514 | 1.38 | 0.528 | 3.59 | 0.509 |
|  |  |  | BG | 0.68 | 1.3 | 0.209 | 1.95 | 0.689 | 5.52 | 0.193 |
|  |  |  | DGM | 0.63 | 1.2 | 0.233 | 1.84 | 0.675 | 5.03 | 0.222 |
|  |  |  | GM | 0.35 | 0.7 | 0.481 | 1.4 | 0.545 | 3.62 | 0.477 |
|  |  |  | HC | 0.26 | 0.5 | 0.626 | 1.29 | 0.46 | 3.63 | 0.622 |
|  |  |  | Thalamus | 0.48 | 0.9 | 0.35 | 1.6 | 0.598 | 4.26 | 0.343 |
|  |  |  | NAWM | 0.39 | 0.7 | 0.454 | 1.46 | 0.543 | 3.91 | 0.451 |
|  |  |  | WML | 0.12 | 0.2 | 0.808 | 1.12 | 0.448 | 2.8 | 0.807 |
|  | YR0 EBNA1 antibodies (log) | YR1 to YR5 change in MTsat | Amygdala | -0.16 | -0.9 | 0.382 | 0.71 | 0.333 | 1.52 | 0.378 |
|  |  |  | BG | -0.14 | -0.8 | 0.431 | 0.74 | 0.353 | 1.56 | 0.426 |
|  |  |  | DGM | -0.13 | -0.7 | 0.468 | 0.76 | 0.356 | 1.61 | 0.461 |
|  |  |  | GM | -0.12 | -0.7 | 0.508 | 0.77 | 0.365 | 1.65 | 0.505 |
|  |  |  | HC | -0.17 | -0.9 | 0.393 | 0.71 | 0.316 | 1.57 | 0.382 |
|  |  |  | Thalamus | -0.1 | -0.6 | 0.578 | 0.8 | 0.374 | 1.73 | 0.574 |
|  |  |  | NAWM | -0.15 | -0.8 | 0.417 | 0.73 | 0.345 | 1.55 | 0.416 |
|  |  |  | WML | -0.13 | -0.7 | 0.503 | 0.76 | 0.346 | 1.68 | 0.494 |
|  |  | YR1 to YR5 change in T1 | Amygdala | -0.06 | -0.3 | 0.746 | 0.88 | 0.407 | 1.91 | 0.746 |
|  |  |  | BG | -0.13 | -0.7 | 0.516 | 0.77 | 0.349 | 1.7 | 0.506 |
|  |  |  | DGM | -0.12 | -0.6 | 0.524 | 0.78 | 0.362 | 1.68 | 0.518 |
|  |  |  | GM | -0.14 | -0.8 | 0.441 | 0.75 | 0.356 | 1.57 | 0.446 |
|  |  |  | HC | -0.09 | -0.4 | 0.656 | 0.83 | 0.376 | 1.85 | 0.654 |
|  |  |  | Thalamus | -0.11 | -0.6 | 0.535 | 0.79 | 0.375 | 1.66 | 0.533 |
|  |  |  | NAWM | -0.13 | -0.7 | 0.459 | 0.76 | 0.361 | 1.58 | 0.461 |
|  |  |  | WML | -0.19 | -1 | 0.325 | 0.67 | 0.308 | 1.48 | 0.313 |
|  | YR0 NfL z-score | YR1 to YR5 change in MTsat | Amygdala | -0.18 | -0.6 | 0.571 | 0.76 | 0.292 | 1.97 | 0.57 |
|  |  |  | BG | -0.19 | -0.6 | 0.554 | 0.74 | 0.275 | 2 | 0.552 |
|  |  |  | DGM | -0.18 | -0.6 | 0.577 | 0.75 | 0.279 | 2.04 | 0.575 |
|  |  |  | GM | -0.17 | -0.5 | 0.607 | 0.77 | 0.282 | 2.1 | 0.605 |
|  |  |  | HC | -0.09 | -0.3 | 0.765 | 0.86 | 0.326 | 2.28 | 0.765 |
|  |  |  | Thalamus | -0.12 | -0.4 | 0.71 | 0.82 | 0.292 | 2.31 | 0.709 |
|  |  |  | NAWM | -0.08 | -0.3 | 0.794 | 0.88 | 0.324 | 2.37 | 0.794 |
|  |  |  | WML | -0.09 | -0.3 | 0.779 | 0.87 | 0.342 | 2.23 | 0.779 |
|  |  | YR1 to YR5 change in T1 | Amygdala | 0 | 0 | 0.999 | 1 | 0.36 | 2.77 | 0.999 |
|  |  |  | BG | 0.03 | 0.1 | 0.929 | 1.05 | 0.369 | 2.98 | 0.929 |
|  |  |  | DGM | 0.01 | 0 | 0.984 | 1.01 | 0.367 | 2.78 | 0.984 |
|  |  |  | GM | -0.1 | -0.3 | 0.745 | 0.85 | 0.325 | 2.23 | 0.743 |
|  |  |  | HC | 0.04 | 0.1 | 0.918 | 1.07 | 0.322 | 3.53 | 0.918 |
|  |  |  | Thalamus | -0.03 | -0.1 | 0.914 | 0.95 | 0.354 | 2.53 | 0.914 |
|  |  |  | NAWM | -0.12 | -0.4 | 0.707 | 0.83 | 0.318 | 2.17 | 0.706 |
|  |  |  | WML | -0.11 | -0.3 | 0.732 | 0.85 | 0.329 | 2.19 | 0.731 |
| *WML: white matter lesions; NAWM: normal-appearing white matter; GM: grey matter; DGM: deep grey matter; HC: hippocampus; BG: basal ganglia; FDR: false discovery rate; ROI: region of interest; OR: odds ratio; YR0: Baseline (year 0); YR1: 1-year follow-up; YR5: 5-year follow-up; MTsat: magnetisation transfer saturation; EDSS: Expanded Disability Status Scale.* | | | | | | | | | | |
